# Physical activity, metabolites, and breast cancer associations

**DOI:** 10.1101/2024.05.10.24307198

**Authors:** Eleanor L. Watts, Steven C. Moore, Leila Abar, Hyokyoung G. Hong, Pedro F. Saint-Maurice, Caitlin O’Connell, Charles E. Matthews, Erikka Loftfield

**Author notes:** Equal contribution (Charles E. Matthews, Erikka Loftfield).

## Abstract

**Background:** The effects of habitual physical activity on physiology and disease prevention are not fully understood. We examined the associations between physical activity, metabolites in systemic circulation, and breast cancer risk.

**Methods:** Total physical activity levels were assessed using doubly labeled water, accelerometers, and previous day recalls in the IDATA study (N=707 participants, ages 50-74 years, 51% women). Assessments occurred 1-6 times over a 12-month period and blood samples were collected twice. Partial Spearman correlations were used to estimate associations between physical activity and 843 serum metabolites, corrected for multiple testing using the false discovery rate (p-adj<0.05). Associations between physical activity-associated metabolites and breast cancer were explored in a prospective cohort (621 cases, 621 controls) using conditional logistic regression.

**Results:** Physical activity was associated with 164 metabolites, spanning a wide range of pathways, including many amino acid pathways, glucose homeostasis, and bile acid metabolism. Nine physical activity-associated metabolites were also associated with postmenopausal breast cancer risk. Key metabolites were N-acetylthreonine, isovalerylglycine, 2-methylbutyroylcarnitine (amino acids and derivatives), androsteroid monosulfate C19H28O6S (1), and X-21310. These metabolites were consistent with a protective role of physical activity on breast cancer prevention and particularly implicated a role for branched chain amino acid catabolism. Sphingomyelin (d18:1/20:1, d18:2/20:0) levels were lower in participants with higher physical activity energy expenditure and were also associated with lower breast cancer risk.

**Conclusion:** Physical activity is associated with a broad range of metabolites, some of which are also associated with reduced breast cancer risk, highlighting potential metabolic pathways for cancer prevention.

## Introduction

Higher doses of physical activity are associated with a lower risk of thirteen cancers including breast cancer[1, 2]. This protective effect is mediated through several key biological mechanisms: weight maintenance, lower sex steroid hormones, reduced low-grade inflammation, and improved immune function and insulin sensitivity[2-4]. The impact of physical activity on other metabolic factors, such as carbohydrate, amino acid, and lipid metabolism, are also likely to be important for cancer prevention[4-6], but have not been extensively explored. Metabolomics, the detailed characterization of many small molecule metabolites, presents an opportunity to systematically explore the complex biological underpinnings of physical activity and its role in disease prevention.

Physical activity, defined as bodily movement by skeletal muscles resulting in energy expenditure, encompasses multiple dimensions including energy used, movements, and specific types of activities[7]. Studies to date of physical activity and metabolite associations have generally highlighted relationships for lipoproteins and amino acids, particularly branched chain amino acids (BCAAs)[8-10]. However, the limited scope of physical activity and metabolites measured have restricted our understanding. For example, the doubly labeled water (DLW) method, considered the gold-standard for measuring energy expenditure under free-living conditions[11], has been explored in one prior study that analyzed 17 metabolic syndrome-associated metabolites in 82 participants[12]. Accelerometers, devices that capture movement, movement intensity, and activity type (e.g., stepping), are commonly used in large-scale epidemiological studies. Higher accelerometer-measured activity is consistently associated with lower breast cancer risk[13]. The range of metabolites investigated in studies exploring the relationships between accelerometer-measured activity and the metabolome is also limited. The most extensive study to date analyzed 328 metabolites, highlighting associations with BCAA pathways and carbohydrate metabolism[10]. No study has investigated the relationship between step counts and metabolites, despite increasing recognition of step count as an important indicator of total activity relevant to cancer[13, 14].

We investigated the associations of 836 serum metabolites with physical activity using DLW, two different accelerometers, and previous day recalls in 700 participants from the Interactive Diet and Activity Tracking in AARP (IDATA) Study. We also explored the associations of the physical activity associated metabolites with breast cancer risk in a nested case control study in the Prostate, Lung, Colorectal and Ovarian (PLCO) Cancer Screening Trial.

## Methods

### The IDATA Study

#### Study design

The IDATA study was designed to evaluate various diet and physical activity measures for their suitability in epidemiologic research. Participants comprised a sample of AARP members (age 50–74 years) from Pittsburgh, Pennsylvania who spoke English, had internet access, were not on a weight-loss diet, had a body mass index (BMI) <40 kg/m^2^, and the absence of major medical conditions and mobility limitations. The study was approved by the NCI Special Studies Institutional Review Board and all participants signed informed consent.

Participants attended the study center up to three times over 12 months between 2012-2013 and completed several diet and physical activity measurements. Participant characteristics, including age and race were obtained from the telephone screen administered prior to the first clinic visit. Body fat mass was measured using deuterium dilution at the DLW assessment; height and weight was measured at each clinical visit.

Smoking information was not directly collected, therefore current smoking intensity was estimated using cotinine, a biological marker of recent smoking[15]. A study timeline for each assessment is available from **Supplementary Table 1**.

A total of 1,082 participants attended the IDATA clinical assessment center and provided specimen consent. We excluded 364 participants who did not have serum metabolite measurements, of which 236 did not complete all biospecimen collections, and 11 participants with missing covariate data (physical activity and age) (**Supplementary Figure 1**).

#### Physical activity assessments

##### DLW

Total energy expenditure (TEE) was measured via DLW for 14 days[16], following established protocols[17, 18]. Resting energy expenditure (REE_pred_) was estimated using the Mifflin-St Jeor equation[19]. To minimize the influence of body size on activity measures, we calculated the physical activity level metric (PAL)=TEE/REE_pred_[11]. Within our analytic sample, 556 participants had one PAL measurement and 29 had two measurements.

##### Accelerometers

Two accelerometers were used to assess overall activity: activPAL 3D and ActiGraph (model GT3X). Both accelerometers were worn twice for 7 days, 6 months apart.

1. activPAL: measured step count, the device was worn on the mid-right thigh continuously (24-hr). Participants recorded the date/time they got out of bed in the morning and into bed each night. Overall, 707 participants had one step count measure and 582 had both measures.
2. ActiGraph: measured physically active metabolic equivalent of task (aMET) hrs/day, using the Sojourn 3x hybrid machine-learning method which estimates intensity via measured energy expenditure in about 30 different activities[20], and we excluded contributions from sedentary time. The device was worn on the right hip from time out of bed until bed for the night. On/off logs were used to estimate the waking day[21]. Overall, 704 participants had one measure of aMET and 613 had both measures.

##### Questionnaire

ACT24 is an internet based previous-day recall instrument, which asks participants to report their time use the previous day using a list of >200 individual activities, including follow-up questions that specify the posture associated with each activity[21]. MET hours/day estimates were derived from linkage with the Physical Activity Compendium[21]. Participants were invited by email to complete up to six ACT24s over the study duration; aMET hrs/day were calculated based on time allocation to each non-sedentary activity. Overall, 706 completed at least one questionnaire and 493 completed all six.

#### Metabolite assessment

For all participants, blood samples were drawn twice six months apart. Blood was cooled, and centrifuged within two hours of collection and stored at -70°C. Metabolites were measured by Metabolon Inc., using ultra-high-performance liquid chromatography with tandem mass spectrometry to separate compounds and measure spectral peaks[22]. Values below the limit of detection were assigned the minimum observed value for each metabolite and normalized by run day. Metabolites levels were log-transformed and standardized to mean 0, standard deviation 1. Metabolites with values below the limit of detection in >80% of participants across the two visits (N=172) and metabolites with a coefficient of variation >25% (N=456) were excluded, leaving a total of 836 metabolites included in the analysis. The median intraclass correlation (ICC) for technical reliability was 0.9. Median temporal ICCs were 0.74, based on linear mixed-effects model to estimate the variance across all measurements[23]. Metabolite descriptors, unique identifiers and Metabolon-assigned pathways are available from the **Supplementary Data**.

##### Population for evaluating breast cancer associations

The Prostate, Lung, Colorectal, and Ovarian Cancer Screening Trial (PLCO) study is a population-based multicenter randomized screening trial of people aged 55-74 years at baseline with no history of prostate, lung, colorectal, or ovarian cancer[24, 25]. This study was approved by institutional review boards at the US National Cancer Institute and the 10 study centers.

Participants were selected from a previous nested case-control study of 621 incident invasive primary breast cancer cases (ICD-9 174.0-174.9) in women who were not using hormone therapy at study baseline or who had an estrogen receptor and/or progesterone receptor negative status[26]. Cases were matched to controls using incidence density sampling based on age at randomization, date of blood collection, and menopausal hormone therapy use. All controls were alive and had no history of cancer at the date of diagnosis for the matched case. The median time from blood collection to breast cancer diagnosis was 6.7 years[26].

At study baseline, participants completed questionnaires to report their smoking, physical activity, weight, and height. Serum samples were collected at the first follow-up visit, approximately one-year after baseline. Metabolites were quantified by Metabolon, using the procedures described above, and the median ICC was 0.94[26].

### Statistical methods

#### I. Physical activity and metabolites

We calculated mean physical activity measures, if ≥2 measurements were available, and mean metabolite levels for all participants. The associations between metabolites and physical activity were estimated using Spearman’s partial correlation, adjusted for age at study enrollment, sex, race, smoking status, and body fat index (body fat (kg)/height(m)^2^). Confidence intervals were estimated by bootstrap (1,000 samples with replacement). Body fat measurements were missing for 150 participants, who did not have DLW measures; this data was imputed using multiple imputation by chained equations (MICE), based on anthropometric measures, age, race, and sex. Multiple testing was controlled for using the false discovery rate within each physical activity measure using the Benjamini-Hochberg procedure (p-adjusted<0.05).

To explore the mechanistic pathways influenced by physical activity, we combined p-values from partial correlation models across pathways using Fisher’s method. This method relies on a null distribution generated from pseudo replicates to estimate the variance-covariance matrix of the test statistics, based on the correlation matrix of relevant metabolites[27]. These pathways, defined by Metabolon, have the limitation of assigning each metabolite to only one pathway. To address this and achieve a broader analysis, we also employed clustered pathway enrichment through RaMP, which integrates annotations from various metabolomic databases and conducts overrepresentation analysis via Fisher’s test[28].

##### Further analysis

We performed subgroup analyses for each metabolite by sex, and tested for heterogeneity in the associations using Cochran’s Q[28], using the p-value set at the same level that was the statistic for significance in the primary analysis (p-unadjusted<0.005). To summarize overall agreement, we calculated Spearman’s pairwise correlation of the physical activity-metabolite correlations between each group (i.e., the correlation of correlations).

The calculation of PAL includes predicted REE, which may lead to a small amount of measurement error[29]. Therefore, we also estimated associations of TEE from DLW with additional adjustment for total fat mass (kg) and fat-free mass (kg) estimated using deuterium dilution.

#### II. Exploratory analysis of physical activity-associated metabolites and breast cancer risk in serum

We explored the associations of each physical activity-associated metabolite with breast cancer risk in the PLCO cohort. Odds ratios (OR) were estimated using conditional logistic regression, conditioned on the matching factors, and adjusted for age at blood draw, age at menarche, age at first live birth and number of live births, type of menopause and age at menopause, menopausal hormone therapy, history of benign breast disease, first-degree family history of breast cancer, race/ethnicity, education, smoking history, diabetes history, Healthy Eating Index 2015 score, alcohol consumption and BMI[26, 30]. Missing covariate data was imputed using MICE, for a small quantity of missing data (≤8%, **Supplementary Table 2**). To identify the key metabolites in the physical activity-breast cancer relationship, we used bidirectional stepwise selection, based on the Akaike Information Criterion[31, 32].

## Results

Participants with higher PAL were younger, had lower body fat index, and were more likely to be female (**Table 1**). Measures of overall physical activity were positively correlated. For instance, PAL was positively correlated with step count, aMET hrs/day (ActiGraph) and aMET hrs/day (ACT24) (r=0.47, 0.53, and 0.36, respectively) (**Supplementary Figure 2**).

**Table 1:** Participant characteristics by tertiles of physical activity level assessed by doubly labelled water.

| Variable | Physical activity level, doubly labeled water |  |  |  |  |  |
| --- | --- | --- | --- | --- | --- | --- |
|  | Tertile 1 (N=186) |  | Tertile 2 (N=185) |  | Tertile 3 (N=185) |  |
|  | % | Mean (SD) | % | Mean (SD) | % | Mean (SD) |
| Age, years |  | 64 (5.9) |  | 63 (5.8) |  | 63 (6.1) |
| Sex |  |  |  |  |  |  |
| Male | 54% |  | 52% |  | 43% |  |
| Female | 46% |  | 48% |  | 57% |  |
| Race |  |  |  |  |  |  |
| Non-Hispanic White | 92% |  | 96% |  | 96% |  |
| Other | 8% |  | 4% |  | 4% |  |
| Height (cm) |  | 171 (8.8) |  | 170 (9.0) |  | 167 (9.2) |
| Weight (kg) |  | 85 (17.0) |  | 81 (16.0) |  | 77 (15.0) |
| BMI (kg/m<sup>2</sup>) |  | 29 (4.8) |  | 28 (4.4) |  | 27 (4.1) |
| Body fat index (body fat/height<sup>2</sup>) |  | 12 (3.6) |  | 11 (3.2) |  | 10 (2.9) |
| Estimated RMR (kcal/day) |  | 1,534 (254) |  | 1,492 (254) |  | 1,412 (255) |
| Doubly labeled water |  |  |  |  |  |  |
| Total activity expenditure (kcal/day) |  | 2,199 (395) |  | 2,466 (423) |  | 2,741 (540) |
| PAEE (kcal/day) |  | 444 (151) |  | 725 (141) |  | 1,055 (279) |
| Physical activity level |  | 1.4 (0.1) |  | 1.7 (0.1) |  | 1.9 (0.2) |
| ActivPAL |  |  |  |  |  |  |
| Steps (n/day) |  | 6,112 (2,313) |  | 7,424 (2,493) |  | 9,207 (3,353) |
| ActiGraph |  |  |  |  |  |  |
| Total MET (hrs/day) |  | 20 (2.7) |  | 22 (2.8) |  | 24 (3.2) |
| Active MET (hrs/day) |  | 11 (3.2) |  | 13 (3.4) |  | 16 (4.1) |
| Sedentary (hrs/day) |  | 8.3 (1.3) |  | 8.2 (1.2) |  | 7.5 (1.4) |
| Light activity (hrs/day) |  | 4.3 (1.2) |  | 4.7 (1.2) |  | 5.4 (1.3) |
| Moderate activity (hrs/day) |  | 0.8 (0.4) |  | 1.0 (0.5) |  | 1.3 (0.6) |
| Vigorous activity (hrs/day) |  | 0.1 (0.1) |  | 0.1 (0.1) |  | 0.2 (0.2) |
| ACT24 |  |  |  |  |  |  |
| Total MET (hrs/day) |  | 36 (3.3) |  | 37 (4.1) |  | 39 (4.8) |
| Active MET (hrs/day) |  | 14 (5.4) |  | 16 (6.3) |  | 20 (7.0) |

Abbreviations: BMI=body mass index, MET=metabolic equivalent of task, PAEE=Physical activity energy expenditure, RMR=resting metabolic rate, SD=standard deviation

### Association of physical activity with metabolites

#### Physical activity-metabolite associations

We identified 164 serum metabolites associated with one or more of the physical activity measures. aMET hrs/day, assessed by ActiGraph, was associated with the highest number of metabolites (N=85), followed by PAL (DLW, N=72), steps/day (activPAL, N=71), and aMET hrs/day (ACT24, N=5) (**Figure 1 and Supplementary Table 3**). One metabolite, 1-methylnicotinamide, was associated with all four physical activity measures.

**Figure 1:**
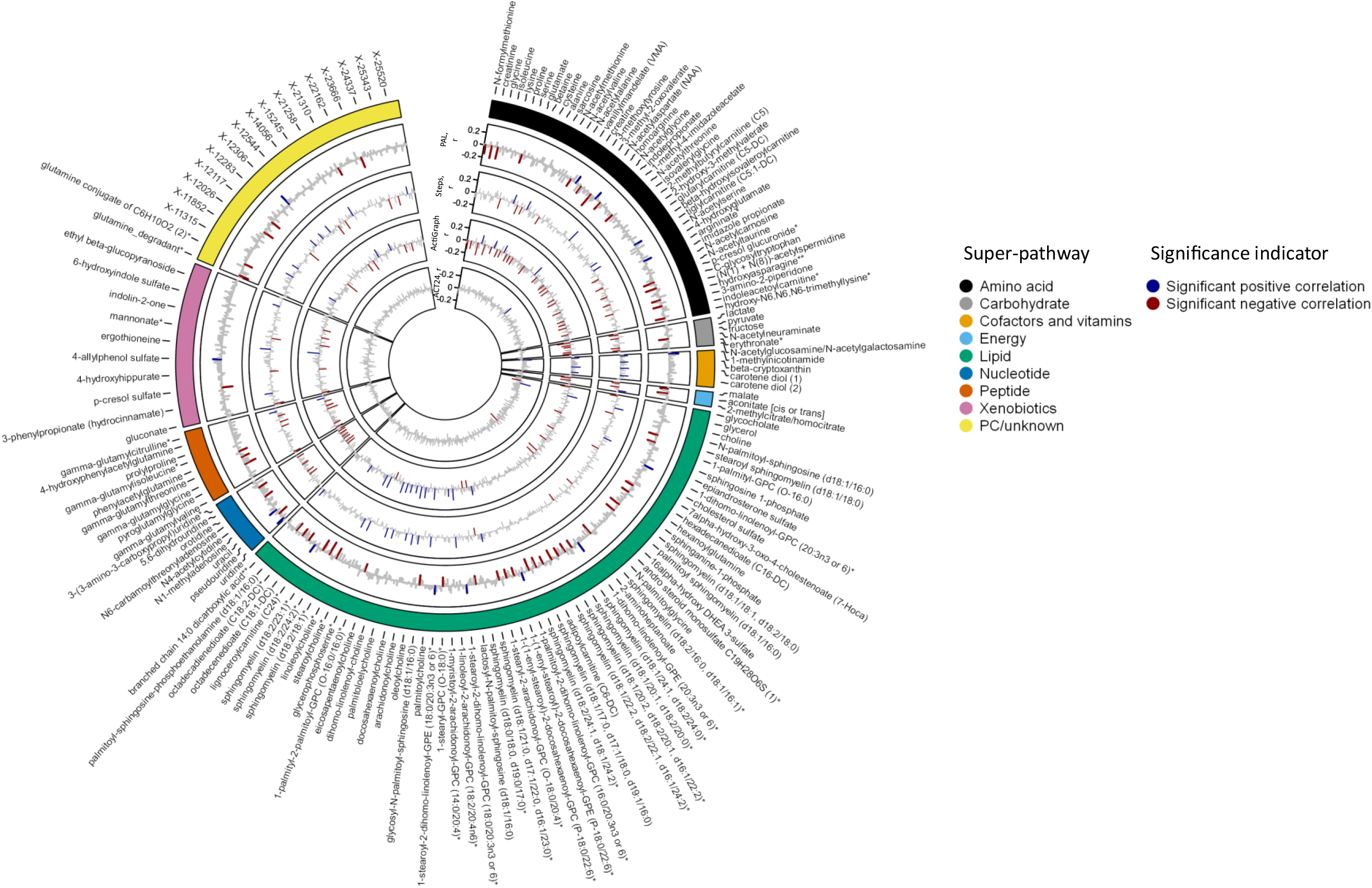
Serum metabolite partial correlations with physical activity. Associations estimated using Spearman partial correlations, adjusted for age (continuous), sex (men, women), smoking (cotinine detected: yes, no), race (non-White, White), body fat index (continuous). Confidence intervals estimated using bootstrap analysis (N iterations=1,000). Significance after correcting for multiple testing using the Benjamini-Hochberg procedure (p-adjusted<0.05). *Indicates a compound that has not been confirmed based on a standard, but confidence in its identity. **Indicates a compound for which a standard is not available, but reasonable confidence in its identity, or the information provided. Metabolon indicator that a compound that has not been confirmed based on a standard, but confidence in its identity. Abbreviations: aMET= active metabolic equivalent of task, PAL=physical activity level, PC=partially characterized.

For PAL, associations were identified for 13 sub-pathways, notably relating to amino acid metabolism, lipid (sphingomyelin and ceramide phosphoethanolamine, and fatty acid metabolism), and pyrimidine metabolism (**Figure 2** and **Supplementary Figure 3**). Most metabolites associated with PAL (80%) were not associated with any other physical activity measure (**Figure 3**). This was most notable for sphingomyelins, which were a distinct and highly correlated group (**Figure 4**).

**Figure 2:**
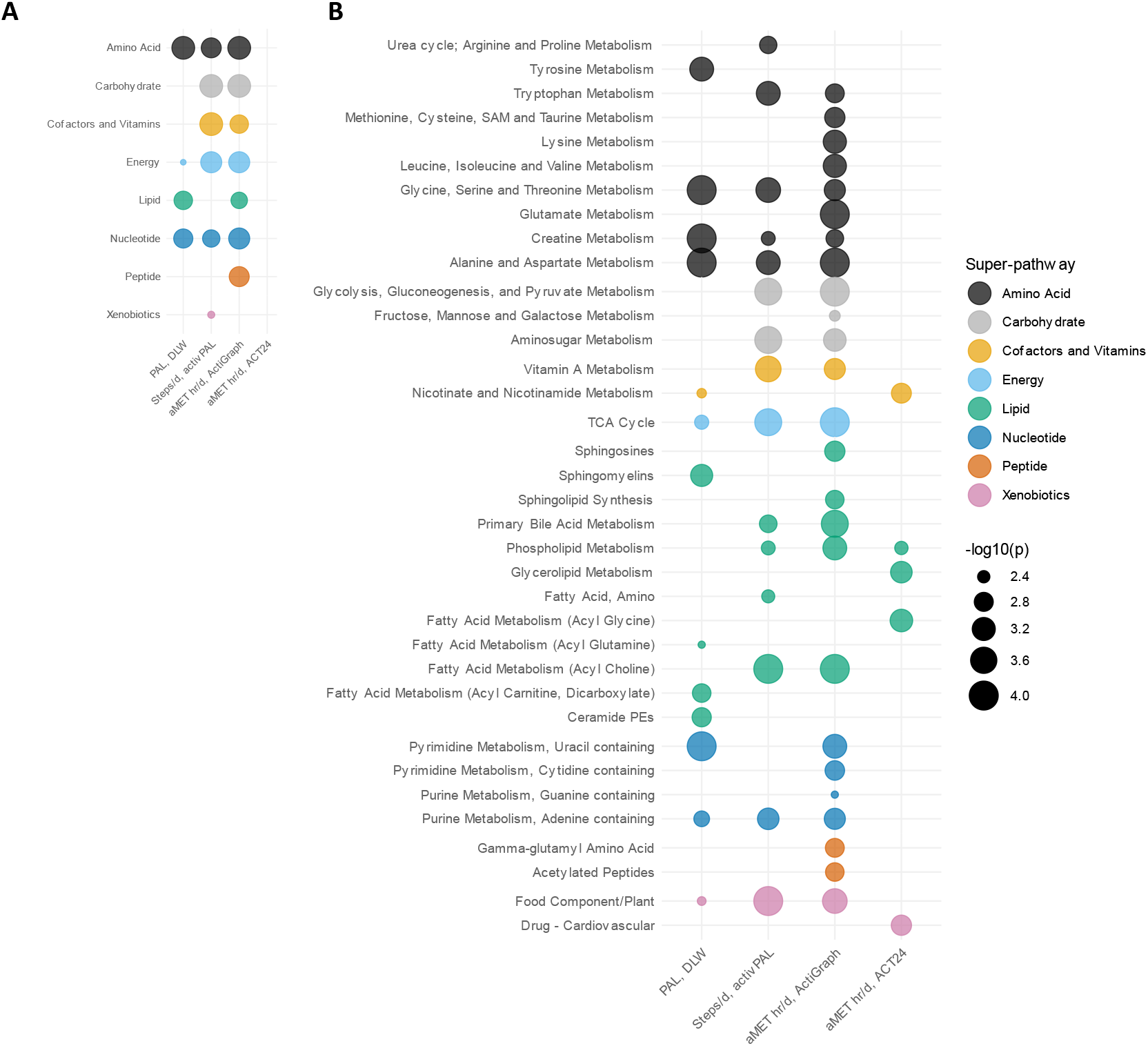
Enrichment analysis of serum metabolites associated with pathways by physical activity measurement. **A) Super-pathway** **B) Sub-pathway** Pathways assigned by Metabolon. P-values derived from Spearman partial correlations, adjusted for age (continuous), sex (men, women), smoking (cotinine detected: yes, no), race (non-White, White), body fat index (continuous) and were pooled by pathway using Fisher’s method. The pathways displayed are for those significantly associated with any of the four measures of physical activity, based on the p-value threshold from the analysis of 836 metabolites, p-value<0.005. Abbreviations: aMET=active metabolic equivalent of task, DLW=doubly labelled water, PAL=physical activity level, PE=phosphoethanolamine, SAM=S-Adenosylmethionine, TCA=tricarboxylic acid.

**Figure 3:**
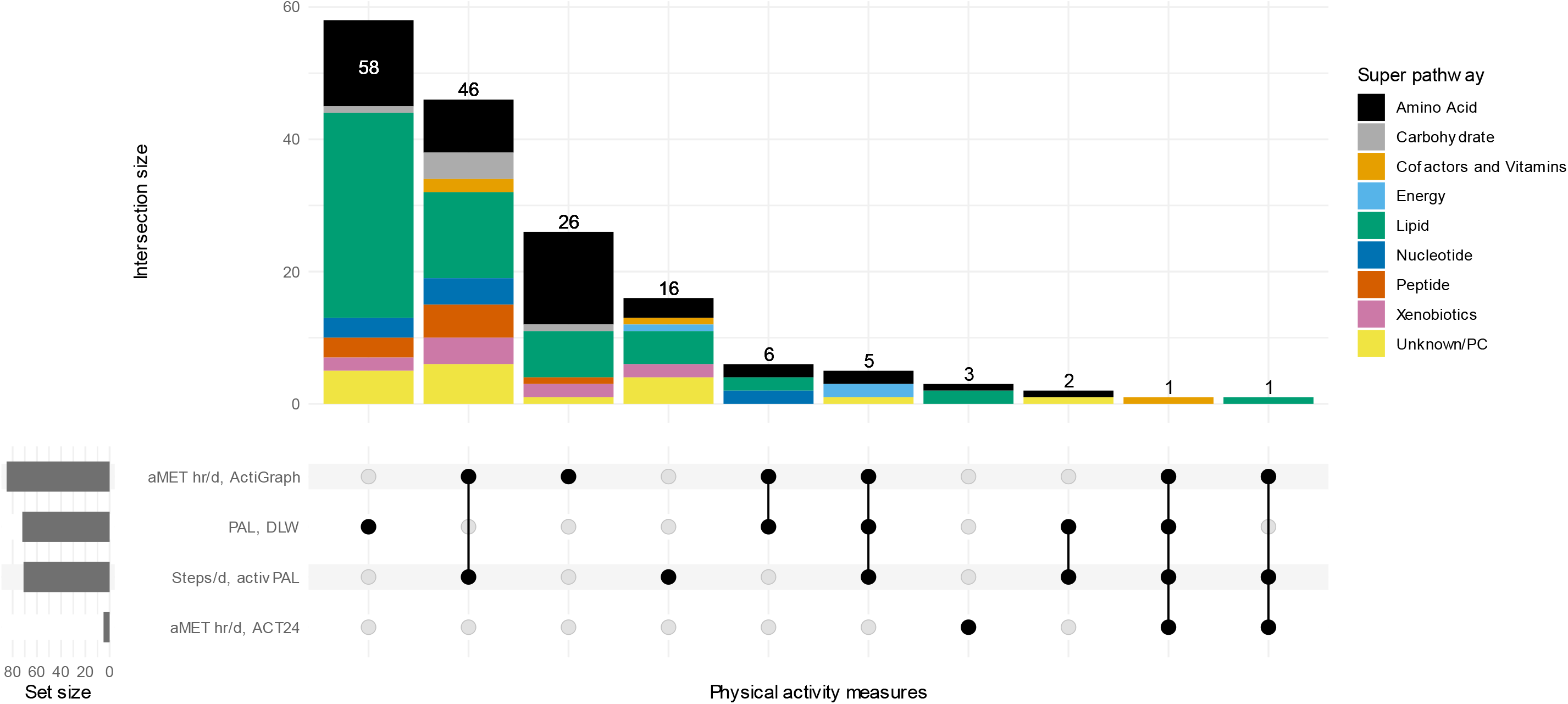
UpSet plot of intersections of serum metabolites associated with physical activity by measure. Correlations estimated using Spearman partial correlation, adjusted for age (continuous), sex (men, women), smoking (cotinine detected: yes, no), race (non-White, White), body fat index (continuous). Multiple testing was corrected for using the Benjamini-Hochberg procedure (p-adjust<0.05). This plot shows the intersections of metabolites associated with different physical activity measures. Vertical bars represent the count of shared metabolites across these measures, with horizontal bars reflecting the total count of metabolites identified by each measure. The matrix at the bottom indicates which physical activity measures are included in each intersection. Abbreviations: aMET=active metabolic equivalent of task, DLW=doubly labelled water, PAL=physical activity level, PC=partially characterized.

**Figure 4:**
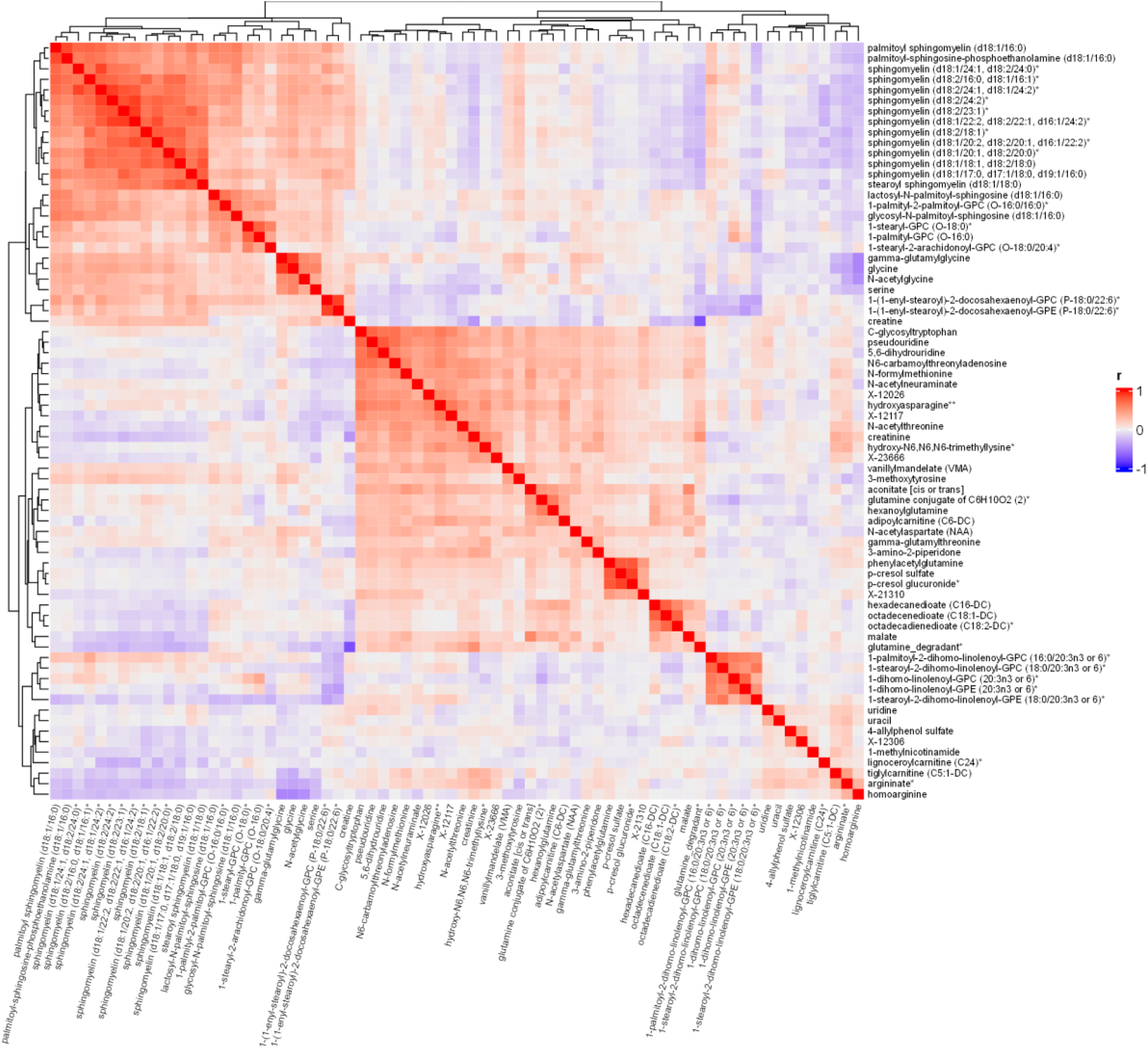
Spearman correlations between serum metabolites associated with physical activity level assessed by doubly labelled water. Metabolites included were associated with physical activity level (assessed by doubly labelled water, based on partial Spearman correlations, adjusted for age (continuous), sex (men, women), smoking (cotinine detected: yes, no), race (non-White, White), body fat index (continuous). Multiple testing was corrected for using the false discovery rate (p-adjusted<0.05). Heatmap created using the ComplexHeatmap R package[54]. *Indicates a compound that has not been confirmed based on a standard, but confidence in its identity. **Indicates a compound for which a standard is not available, but reasonable confidence in its identity, or the information provided.

For step count and aMET hrs/day assessed by accelerometers, 27 sub-pathways were identified, which were highly similar between measures. These pathways encompassed a broad range of functions including amino acid metabolism, glucose homeostasis and tricarboxylic acid (TCA) cycle, as well as bile acid, fatty acid, pyrimidine, and purine metabolism (**Figure 2** and **Supplementary Figures 4-5**). Choline metabolites formed a distinct and highly correlated group across both measures (**Supplementary Figures 6-7**). In total, 46 metabolites were associated with both step count and aMET hrs/day (**Figure 3**) and the overall agreement (i.e., the correlation of correlations) was high (r=0.87) (**Supplementary Figure 8**).

For aMET hrs/day assessed by ACT24, five sub-pathways were identified: nicotinate, and nicotinamide, phospholipid, glycerolipid, fatty acid metabolism (acyl glycine), and cardiovascular drugs (**Figure 2 and Supplementary Figure 9**). Significant metabolites were not strongly correlated with each other (**Supplementary Figure 10**). Agreement between ACT24 and the accelerometers was moderate (r=0.66, **Supplementary Figure 8**).

#### Further analyses

Agreement of metabolite associations by sex was low-to-moderate (agreement=0.21-0.60, **Supplementary Figure 11**) and we observed significant heterogeneity in the associations of four metabolites (**Supplementary Table 4**). Associations for each metabolite by sex are available from **Supplementary Data 2**. The associations of TEE with metabolites, adjusted for fat mass and fat-free mass, were broadly similar to the associations for PAL (**Supplementary Table 5**).

##### Analysis of physical activity associated metabolites in serum with breast cancer

Participants in the nested PLCO study were mean 64 years old, had a mean BMI 27 kg/m^2^ and 90% non-Hispanic White (**Supplementary Table 2**). Of the 164 physical activity-associated metabolites identified in IDATA, 97 were available in our PLCO dataset, and nine were nominally associated with breast cancer risk. Six of these metabolites were consistent with a protective effect of physical activity on breast cancer, of which five metabolites were lower in participants with higher activity levels and were also associated with an increased risk of breast cancer: 2-methylbutyroylcarnitine (OR per 1 SD=1.17, 95% CI 1.03-1.32), N-acetylthreonine (1.18, 1.05-1.33), androsteroid monosulfate C19H28O6S (1) (1.14, 1.00-1.29), N-acetyltaurine (1.15, 1.02-1.29) and X-21310 (1.14, 1.01-1.28). Isovalerylglycine was higher in participants with higher aMET hr/day (ACT24) and was inversely associated with cancer (0.86, 076-0.98). Three metabolites were not consistent with a protective effect. Two metabolites, beta-hydroxyisovaleroylcarnitine and epiandrosterone sulfate, were positively associated with aMET hrs/day (actiGraph) and cancer risk (1.13, 1.01-1.26 and 1.13, 1.00-1.27, respectively). Sphingomyelin (d18:1/20:1, d18:2/20:0) was inversely associated with both PAL and cancer (0.82, 0.73-0.93) (**Figure 5**).

**Figure 5:**
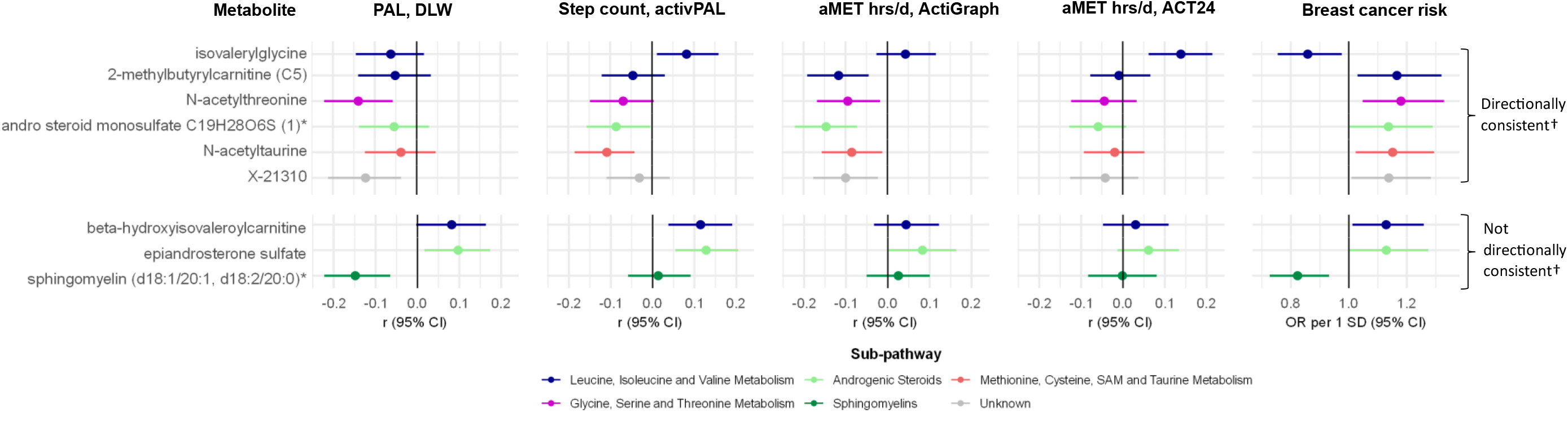
Correlations of metabolites with physical activity measurements in IDATA and their associations with breast cancer risk in PLCO. Associations of metabolites with physical activity were estimated using Spearman partial correlation, adjusted for age (continuous), sex (men, women), smoking (cotinine detected: yes, no), race (non-White, White), body fat index (continuous) in the IDATA study. Confidence intervals estimated using bootstrap analysis (N iterations=1000). Associations with breast cancer risk estimated using conditional logistic regression, conditioned on the matching factors (age at randomization (+/-2 years), date of blood collection (+/-3 months), and menopausal hormone therapy use (current, former/never)), and adjusted for age at blood draw (continuous), age at menarche (<12, 12-13, 14+ years), age at first live birth and number of live births (No live births, <20 years, 20-24 years and 1-2 live births, 20-24 years and >2 live births, 25-29 years and 1-2 live births, 25-29 years and >2 live births, 30+ years), type of menopause and age at menopause (natural menopause: <45 years, natural menopause: 45-49 years, natural menopause: 50-54 years, natural menopause: 55+ years, bilateral oophorectomy/surgery, drug therapy/radiation, hysterectomy with no bilateral oophorectomy), menopausal hormone therapy (never, current. Former), history of benign breast disease (yes, no), first-degree family history of breast cancer (yes. no), race/ethnicity (non-Hispanic white, other), education (up to high school, post high school training other than college, some college, college graduate, postgraduate), smoking history (never, former, current), diabetes status (no, yes), healthy eating index (quartiles), alcohol consumption (0, >0-1, >1-2, >2-4, 4+ drinks/day), body mass index (<25, 25-<30, 30+ kg/m^2^). Metabolites shown here are those associated with any of the physical activity measure (after correcting for multiple testing using the Benjamini-Hochberg procedure, p-adjust <0.05) and nominally associated with breast cancer (p<0.05). *Indicates a compound that has not been confirmed based on a standard, but confidence in its identity. †Directionally consistent metabolites are those with opposing associations for physical activity and breast cancer. Abbreviations: aMET=active metabolic equivalent of task, CI=confidence interval, DLW=doubly labelled water, OR=odds ratio, PAL=physical activity level, PLCO=Prostate, Lung, Colorectal and Ovarian Cancer Screening Trial, SD=standard deviation.

Using stepwise logistic regression, N-acetylthreonine, isovalerylglycine, 2-methylbutyroylcarnitine, androsteroid monosulfate C19H28O6S (1), X-21310, and sphingomyelin (d18:1/20:1, d18:2/20:0) were retained in the model (**Table 2**).

**Table 2:**
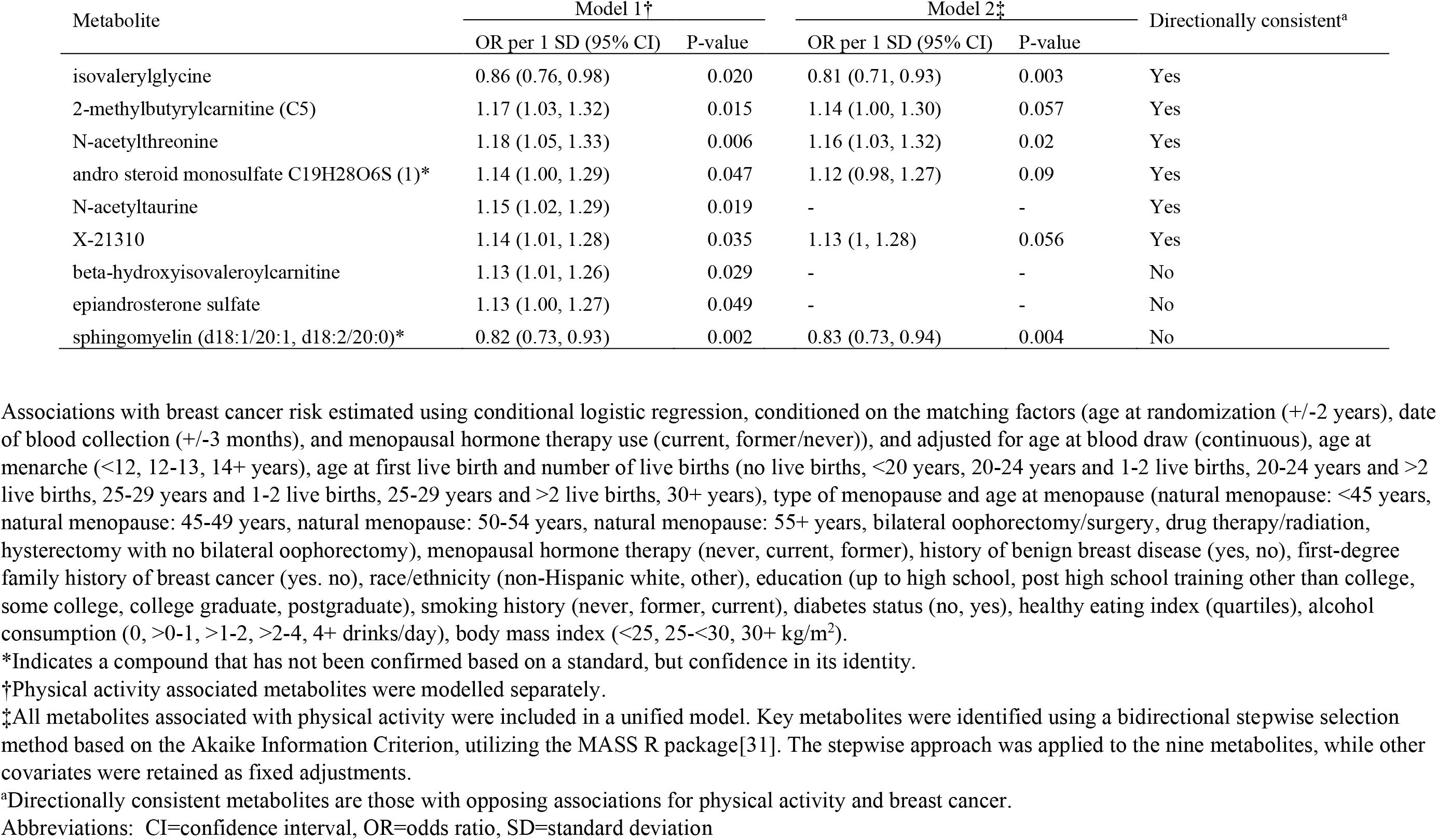
Associations of physical activity associated metabolites with breast cancer risk modelled separately and together using stepwise selection.

| Metabolite | Model 1† |  | Model 2‡ |  | Directionally consistent<sup>a</sup> |
| --- | --- | --- | --- | --- | --- |
|  | OR per 1 SD (95% CI) | P-value | OR per 1 SD (95% CI) | P-value |  |
| isovalerylglycine | 0.86 (0.76, 0.98) | 0.020 | 0.81 (0.71, 0.93) | 0.003 | Yes |
| 2-methylbutyrylcarnitine (C5) | 1.17 (1.03, 1.32) | 0.015 | 1.14 (1.00, 1.30) | 0.057 | Yes |
| N-acetylthreonine | 1.18 (1.05, 1.33) | 0.006 | 1.16 (1.03, 1.32) | 0.02 | Yes |
| andro steroid monosulfate C19H28O6S (1)* | 1.14 (1.00, 1.29) | 0.047 | 1.12 (0.98, 1.27) | 0.09 | Yes |
| N-acetyltaurine | 1.15 (1.02, 1.29) | 0.019 | - | - | Yes |
| X-21310 | 1.14 (1.01, 1.28) | 0.035 | 1.13 (1, 1.28) | 0.056 | Yes |
| beta-hydroxyisovaleroylcarnitine | 1.13 (1.01, 1.26) | 0.029 | - | - | No |
| epiandrosterone sulfate | 1.13 (1.00, 1.27) | 0.049 | - | - | No |
| sphingomyelin (d18:1/20:1, d18:2/20:0)* | 0.82 (0.73, 0.93) | 0.002 | 0.83 (0.73, 0.94) | 0.004 | No |

\*Indicates a compound that has not been confirmed based on a standard, but confidence in its identity.
†Physical activity associated metabolites were modelled separately.
‡All metabolites associated with physical activity were included in a unified model. Key metabolites were identified using a bidirectional stepwise selection method based on the Akaike Information Criterion, utilizing the MASS R package[31]. The stepwise approach was applied to the nine metabolites, while other covariates were retained as fixed adjustments.
<sup>a</sup>Directionally consistent metabolites are those with opposing associations for physical activity and breast cancer.
Abbreviations: CI=confidence interval, OR=odds ratio, SD=standard deviation

The correlations of these nine metabolites with physical activity were not appreciably different between men and women (**Supplementary Table 6**). The associations for each metabolite with breast cancer risk are available from **Supplementary Data 3**.

## Discussion

This is among the most comprehensive study of physical activity and metabolomics available. We identified 164 distinct metabolites associated with activity, stemming from a broad range of biological pathways. Notably, different physical activity measures were associated with different metabolites, possibly suggesting distinct biological effects. In exploratory analyses in a separate prospective study, nine of the physical activity-associated metabolites were also associated with breast cancer risk, with six demonstrating associations that align with the known protective role of physical activity.

Consistent with previous research, we found inverse associations between physical activity and certain amino acids and their derivatives, including isoleucine[9, 10, 33, 34], 3-methyl-2-oxovalerate[10], alanine[34], proline[34], glutamate[10, 34, 35], and creatinine[8]. Additionally, we observed associations with metabolites involved in energy metabolism, protein synthesis and amino acid transport, including pyruvate[33], lactate[34], N1-methyladenosine[36], and gamma-glutamylvaline[10]. In line with prior studies, we observed positive associations with the amino acids and their derivatives, including betaine[10, 34], lysine[12], tiglylcarnitine[35] and with choline[34] and creatine[12]. The wide range of metabolites and physical activity measures in our study also allowed us to identify novel associations, particularly the association of expenditure-based activity, measured by DLW, with lower levels of many sphingomyelin metabolites.

Step count and aMET hrs/day assessed by accelerometer were associated with a broad range of metabolites, though aMET hrs/day identified slightly more metabolites. Strongly implicated pathways include a broad range of amino acid pathways, glucose homeostasis, and fatty acid and bile acid metabolism, and molecular transport systems, which is in line with previous studies[9, 10, 33-38]. However, the breadth of associations across many biological pathways are much wider than previously reported, particularly for energy, nucleotide, and peptide metabolites, likely relating to greater scope of metabolites measured.

We observed notable differences in the associations between physical activity measures and specific metabolites, particularly for PAL. Metabolites associated with PAL, particularly cell membrane lipids such as sphingomyelins, plasmalogens, lysophospholipids, and lysoplasmalogens, were generally not associated with other activity measures. The biological mechanisms underlying these associations remain unclear, but these metabolites may be markers of increased cell turnover associated with higher energy expenditure. Differences between measurements could relate to the distinct dimensions of physical activity each measure captures. For example, DLW measures of energy expenditure are a consequence of bodily movement, but they do not directly capture information about movement associated with distinct behaviors. In contrast, accelerometers, which measure ambulatory activity, might detect short durations of high-intensity activity or be more sensitive to variations in activity patterns. These short durations and variations might contribute minimally to average energy expenditure yet might significantly affect metabolic pathways. We also observed weaker associations for aMET hrs/day, as estimated through up to six days of recall questionnaires. This may relate to the greater intra-individual variability in activity when comparing 6-days of assessment (recalls) vs. 14-days of assessment for DLW and the accelerometers, likely attenuating metabolite associations[21, 39]. These findings underscore the complexity of capturing and interpreting the metabolic associations of physical activity across different activity measures.

We identified nine metabolites associated with both physical activity and breast cancer, three of which are involved in BCAA signaling. Associations with breast cancer appeared to be driven by isovalerylglycine and 2-methylbutyrylcarnitine, byproducts of BCAA catabolism. BCAAs are postulated to affect cancer risk through activation of the mTOR signaling pathway, which regulates cell survival and proliferation[40-43]. Previous studies have reported inconsistent associations between BCAAs and breast cancer, with less understood about their downstream metabolites[44, 45]. Our results suggest that physical activity may modulate BCAA downstream signaling pathways and associate with cancer risk, and we did not observe associations with BCAAs. We also observed associations with androgenic steroids, particularly androsteroid monosulfate C19H28O6S (1). Previous studies have also reported associations between higher physical activity and lower androgen levels[3], and the positive association androgen levels and higher breast cancer risk is well-characterized[46-49]. Conversely, the associations of N-acetylthreonine. X-21310 and sphingomyelin are less understood. Speculatively, the association of N-acetylthreonine might suggest a role for physical activity in modulating acetylation processes (notably 11 of the amino acids associated with physical activity had N-acetyl modifications)[50]. While specific sphingomyelins have been reported to associate with both increased and decreased risks of cardiometabolic diseases and possibly cancer[45, 51-53], these associations warrant further investigation.

The greatest strength of this study is the comprehensive measurements of physical activity, capturing a broad spectrum of activity dimensions with high precision[21]. Physical activity was measured using three validated measures that capture multiple dimensions of physical activity and serum metabolites were measured twice. Therefore, our findings are more likely to reflect usual activity and metabolite relationships, minimizing intra-individual and seasonal variability[39]. Furthermore, deuterium dilution to measure body fat reduces the potential for confounding by adiposity.

Limitations include the observational study design, meaning that we cannot eliminate the possibility of residual confounding both in identifying metabolite associations with physical activity and breast cancer risk, such as from chronic disease. Additionally, 40% of physical activity-associated metabolites were not available in the PLCO breast cancer study, limiting investigation into breast cancer implications. Physical activity was assessed over one year, therefore associations with long-term activity are unknown. Additionally, DLW and ACT24 was administered over different durations than the accelerometers, so it is difficult to directly compare associations. Participants of the IDATA and PLCO studies were >90% non-Hispanic White, which may limit the generalizability of our findings to other populations.

In conclusion, physical activity is associated with a broad range of metabolites and underlying biological pathways, including many amino acid pathways, glucose homeostasis, protein, and fatty acid metabolism. Our findings underscore the potential mechanisms, particularly the modulation of BCAA catabolism, through which physical activity may prevent breast cancer.

## Data Availability Statement

All bona fide researchers can apply to use the IDATA and PLCO resources for health-related research (https://cdas.cancer.gov/).

