## Supplementary Tables and Figures for "Physical activity, metabolites, and breast cancer associations"

**Physical activity, metabolites, and breast cancer associations: Supplementary Tables and Figures**

**Supplementary Figures**

**Supplementary Tables**

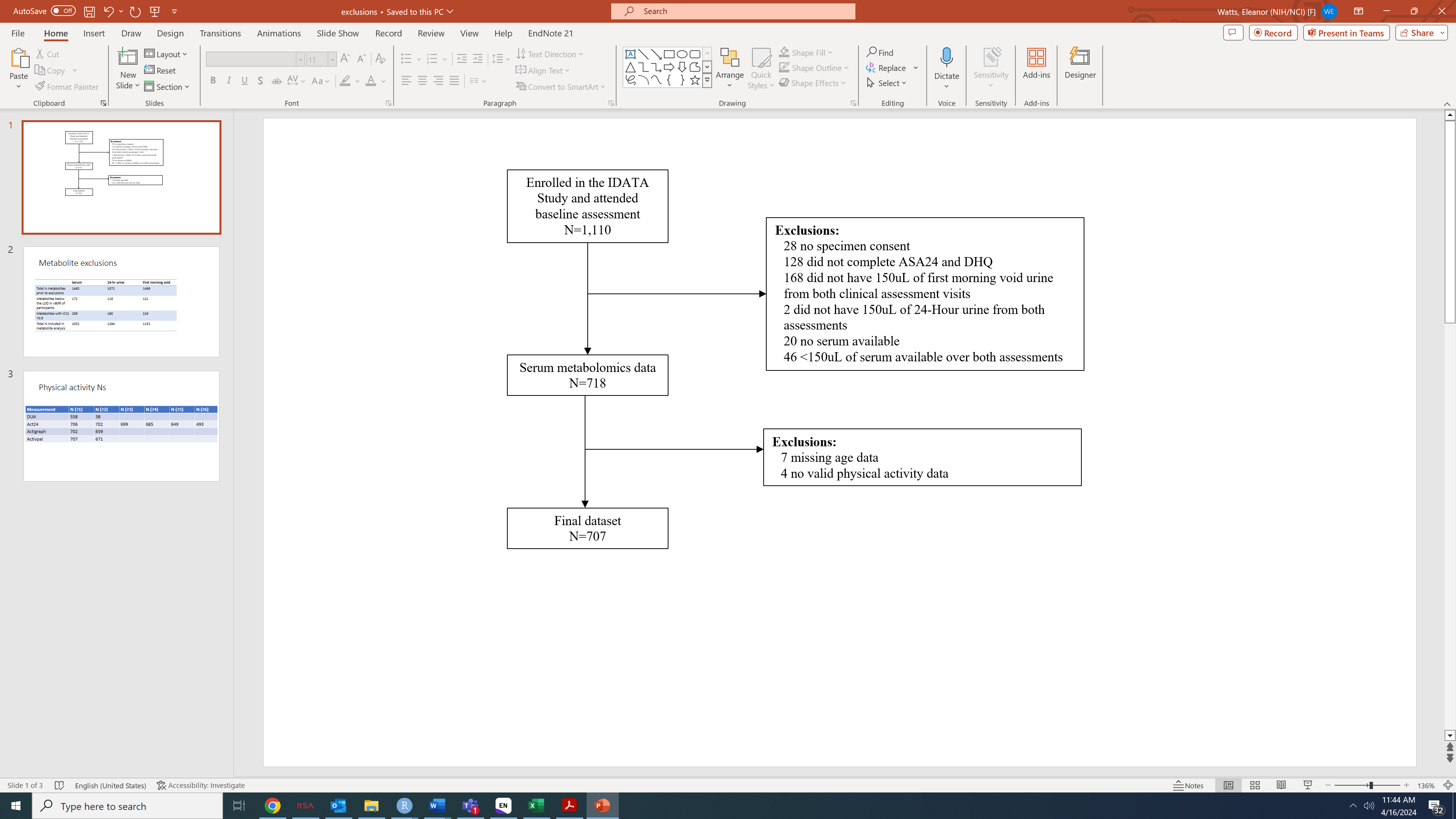

**Supplementary Figure 1: Participant selection flow chart**

Abbreviations: ASA24=Automated Self-Administered 24-Hour, DHQ=Diet History Questionnaire.

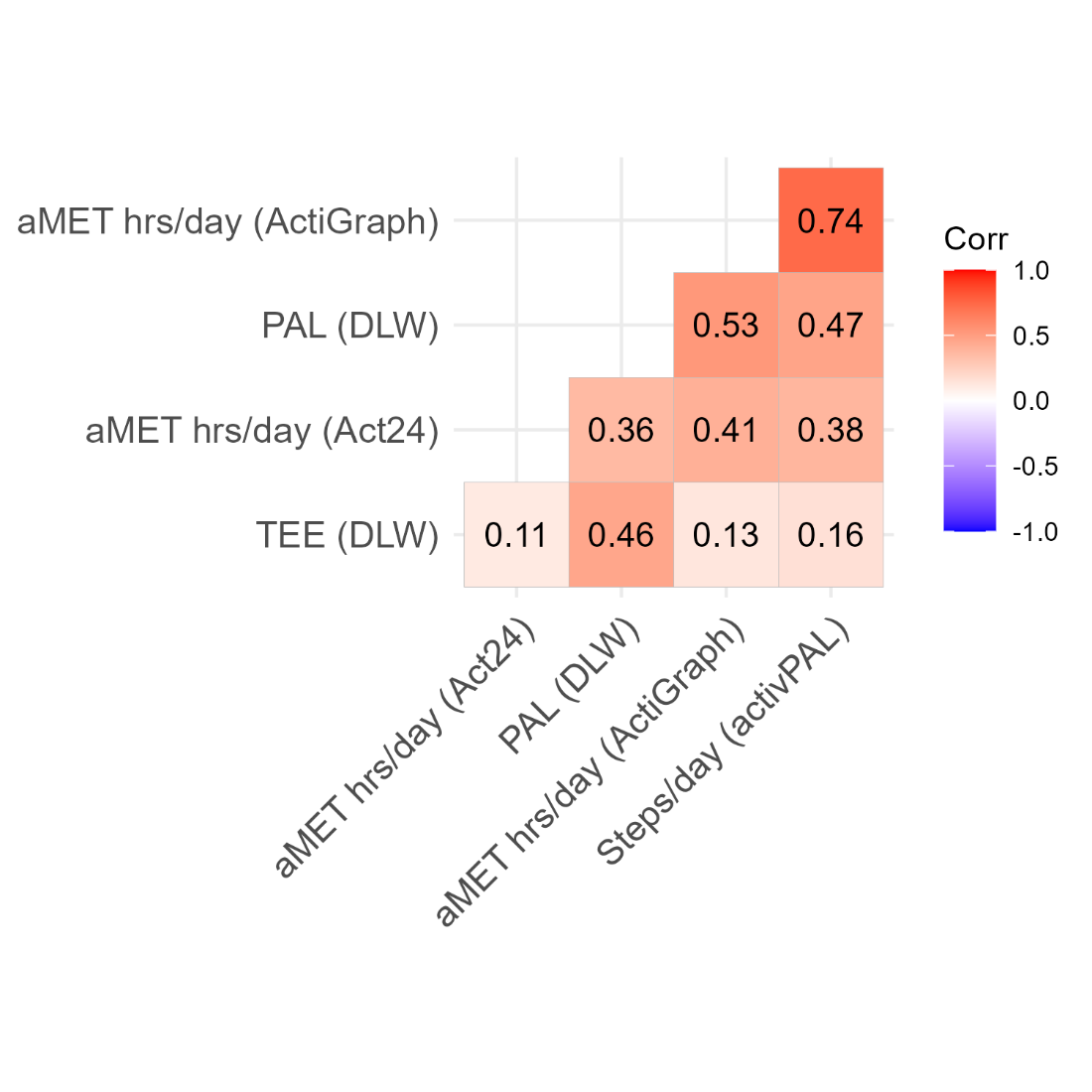

Supplementary Figure 2: Pairwise Spearman correlations between usual physical activity measurements

Abbreviations: aMET= active metabolic equivalent of task, DLW=doubly labelled water, MET=metabolic equivalent of task, PAL=physical activity level, TEE=total energy expenditure.

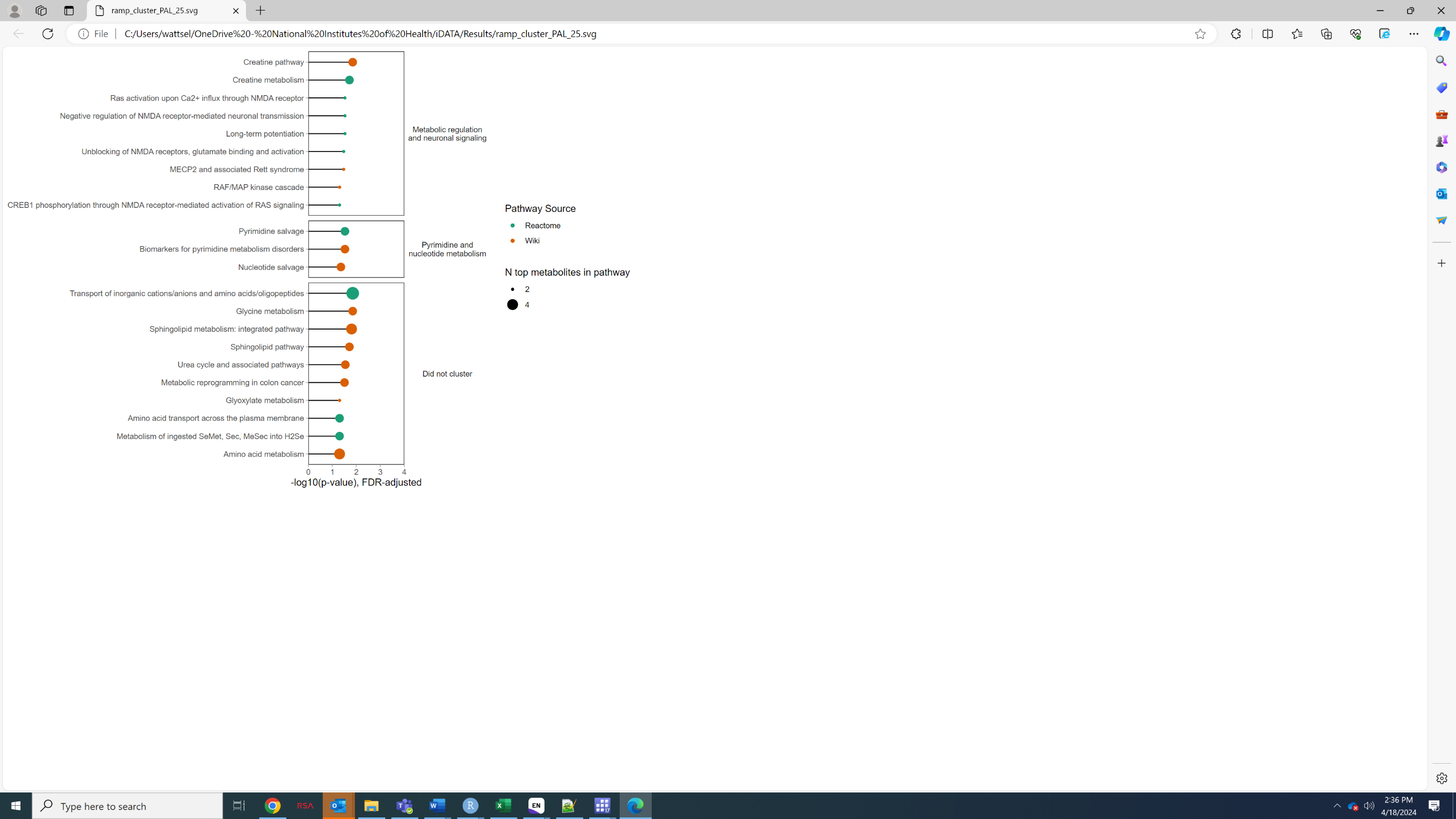

Corrected for multiple testing using the Benjamini-Hochberg Procedure (p-adj<0.05), minimum number of pathways to cluster=3. Minimum overlap for pathways to be considered similar= 20%, minimum overlap for clusters to merge=20%. Clusters annotated to provide summary description.

Supplementary Figure 3: Cluster pathway analysis for metabolites associated with physical activity level, assessed by doubly labelled water using RaMP

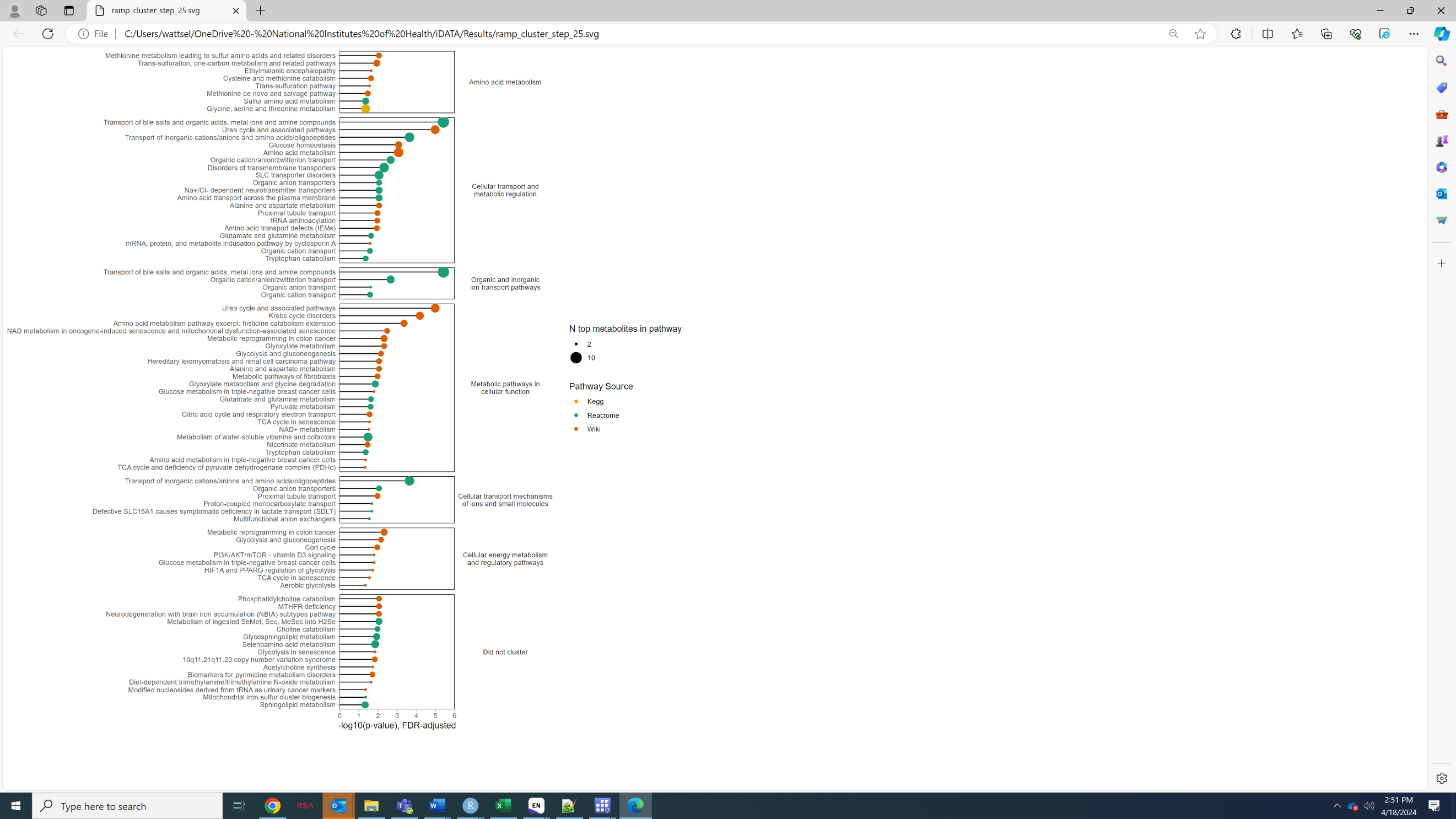

Corrected for multiple testing using the Benjamini-Hochberg Procedure (p-adj<0.05), minimum number of pathways to cluster=3. Minimum overlap for pathways to be considered similar= 20%, minimum overlap for clusters to merge=20%. Clusters annotated to provide summary description.

Supplementary Figure 4: Cluster pathway analysis for metabolites associated with steps/d, assessed by activPAL using RaMP

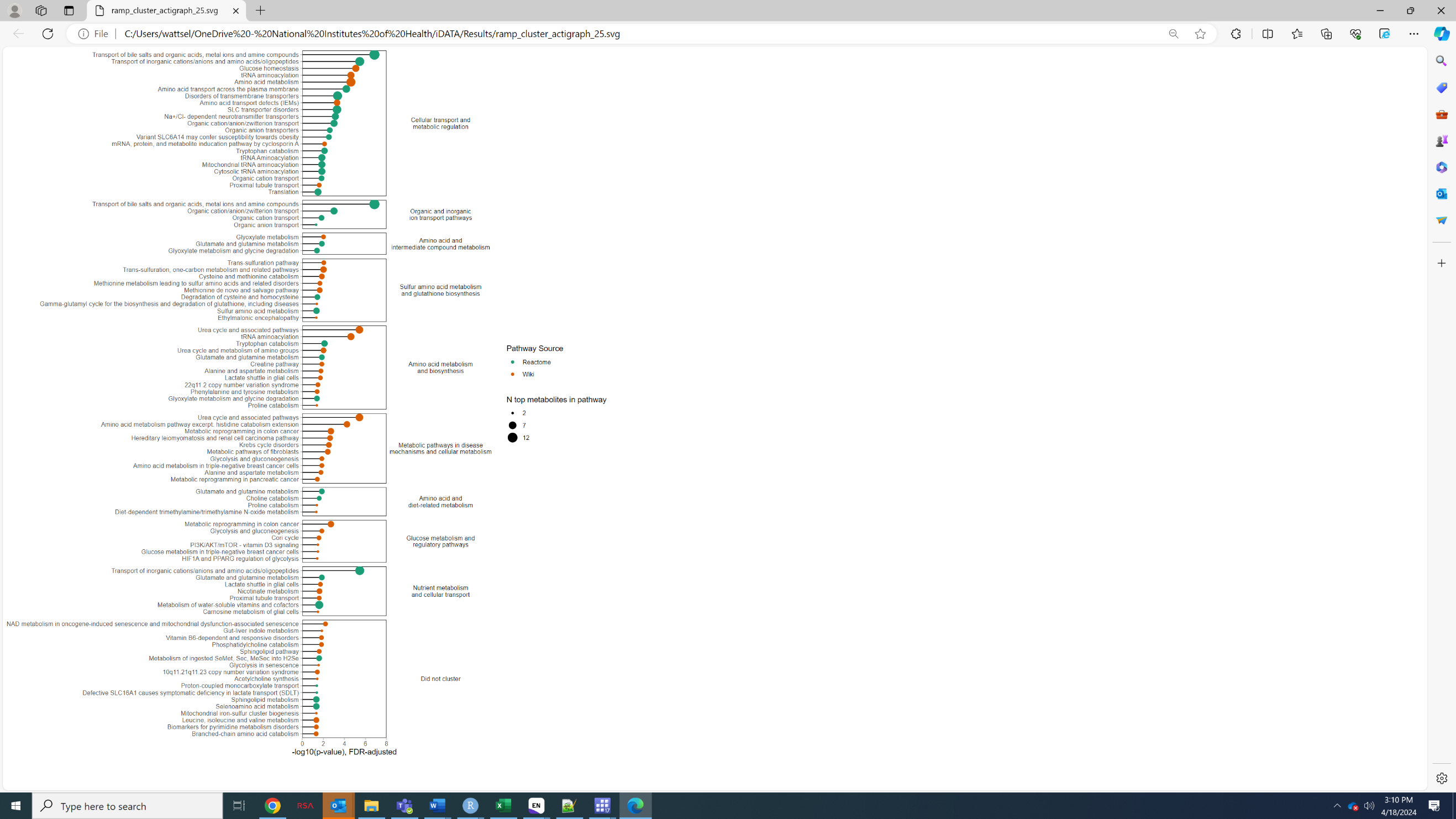

Corrected for multiple testing using the Benjamini-Hochberg Procedure (p-adj<0.05), minimum number of pathways to cluster=3. Minimum overlap for pathways to be considered similar=20%, Minimum overlap for clusters to merge=20%. Clusters annotated to provide summary description.

Abbreviations: aMET=active metabolic equivalent of task

Supplementary Figure 5: Cluster pathway analysis for metabolites associated with aMET hrs/d, assessed by ActiGraph using RaMP

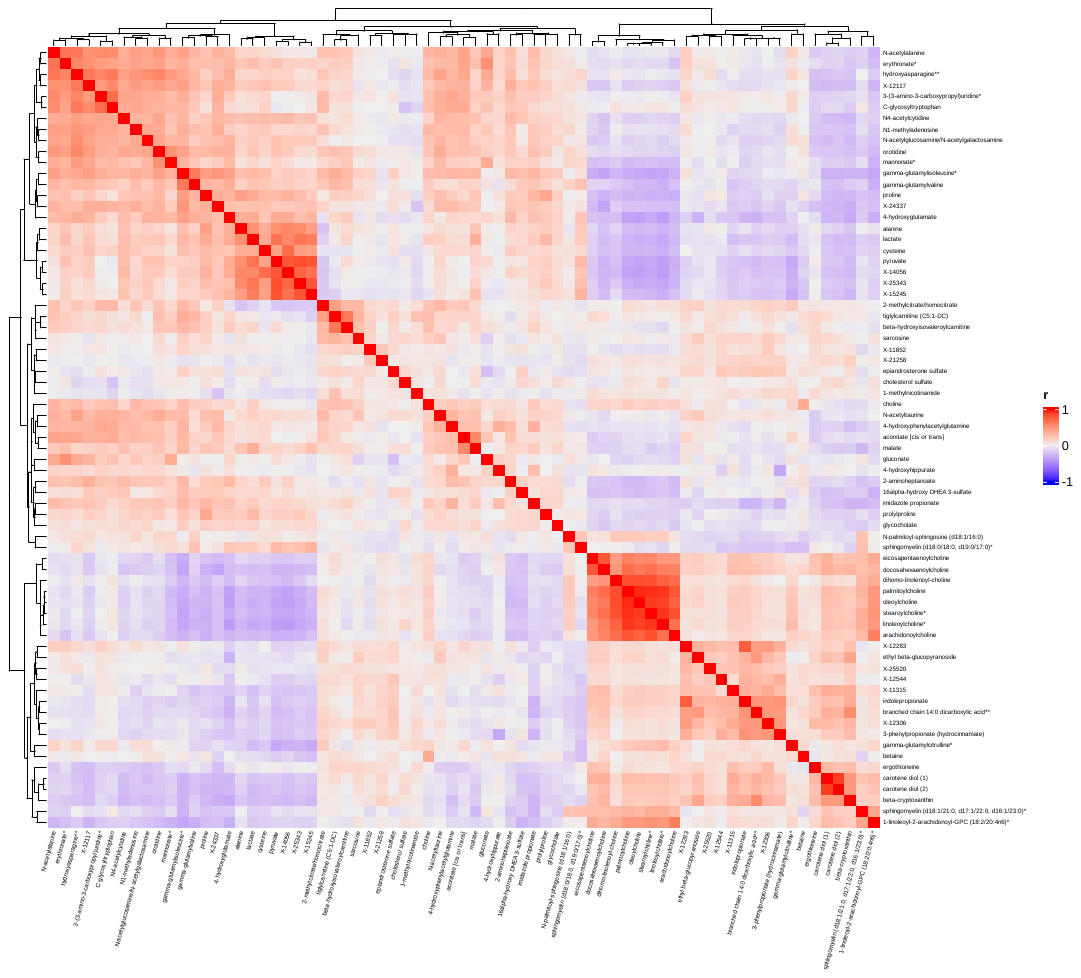

Supplementary Figure 6: Spearman correlations between serum metabolites associated with step count assessed by activPAL

Metabolites included were associated with step count (activPAL) based on partial Spearman correlations, adjusted for age (continuous), sex (men, women), smoking (cotinine detected: yes, no), race (non-White, White), body fat index (continuous). Multiple testing was corrected for using the false discovery rate (p-adjusted<0.05). Heatmap created using the ComplexHeatmap R package^1^**.**

*Indicates a compound that has not been confirmed based on a standard, but confidence in its identity.

**Indicates a compound for which a standard is not available, but reasonable confidence in its identity, or the information provided.

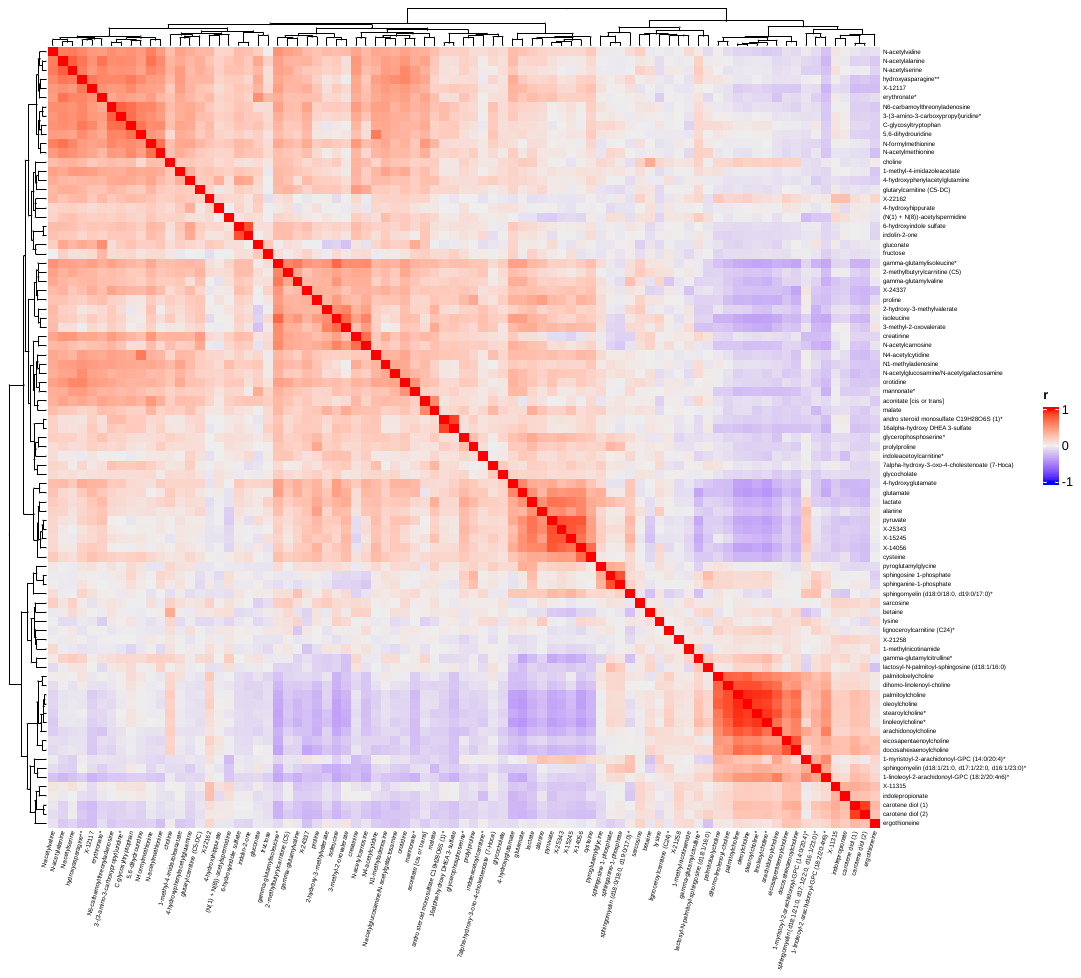

Supplementary Figure 7: Spearman correlations between serum metabolites associated with aMET hrs/day assessed by ActiGraph

Metabolites included were associated with aMET hrs/day (assessed by ActiGraph), based on partial Spearman correlations, adjusted for age (continuous), sex (men, women), smoking (cotinine detected: yes, no), race (non-White, White), body fat index (continuous). Multiple testing was corrected for using the false discovery rate (p-adjusted<0.05). Heatmap created using the ComplexHeatmap R package^1^.

*Indicates a compound that has not been confirmed based on a standard, but confidence in its identity.

**Indicates a compound for which a standard is not available, but reasonable confidence in its identity, or the information provided.

Abbreviations: aMET=active metabolic equivalent of task.

Step count correlation coefficient

Step count correlation coefficient

aMET hrs/d ActiGraph correlation coefficient

aMET hrs/d ActiGraph correlation coefficient

aMET hrs/d ACT24 correlation coefficient

aMET hrs/d ACT24 correlation coefficient

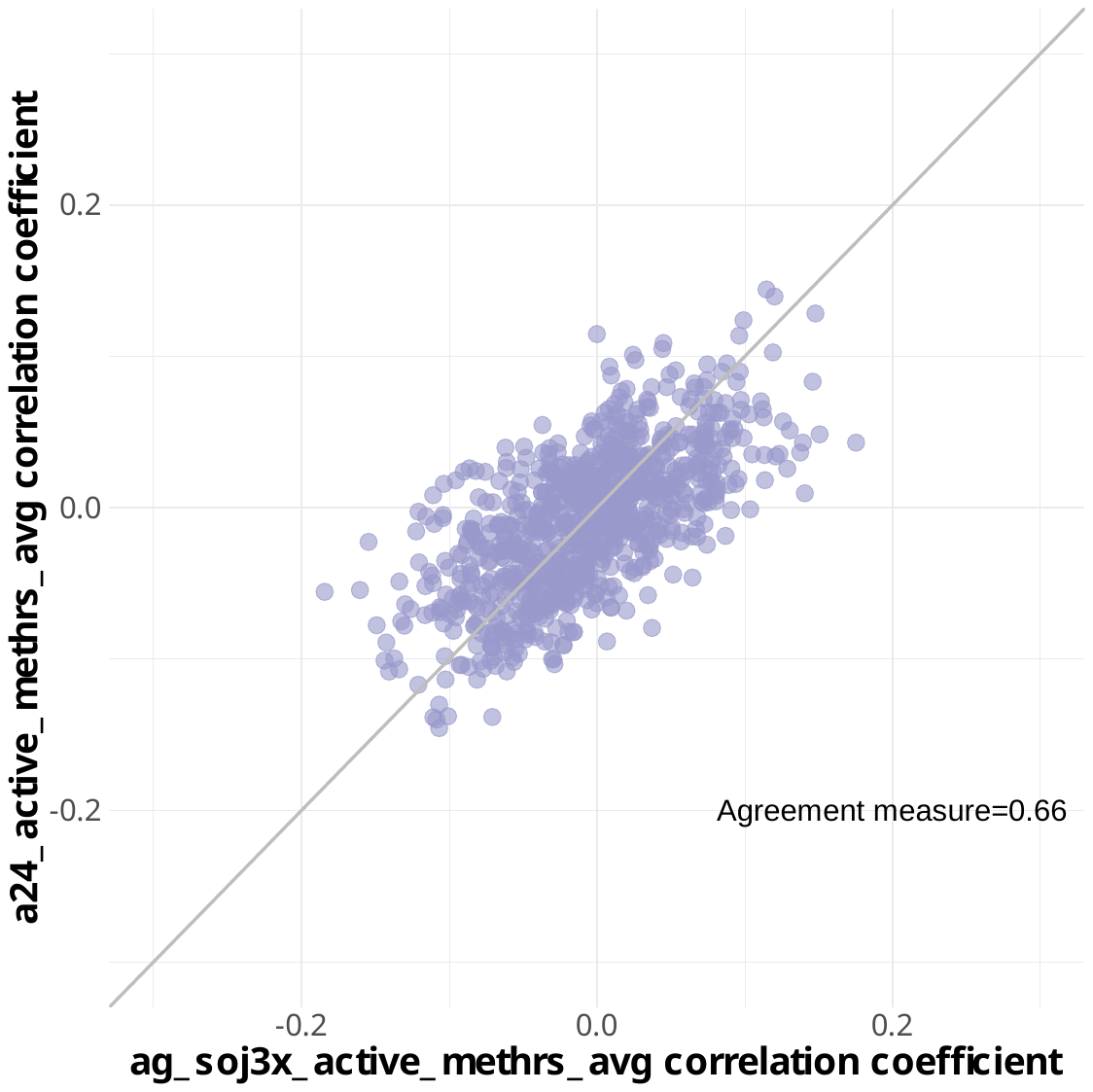

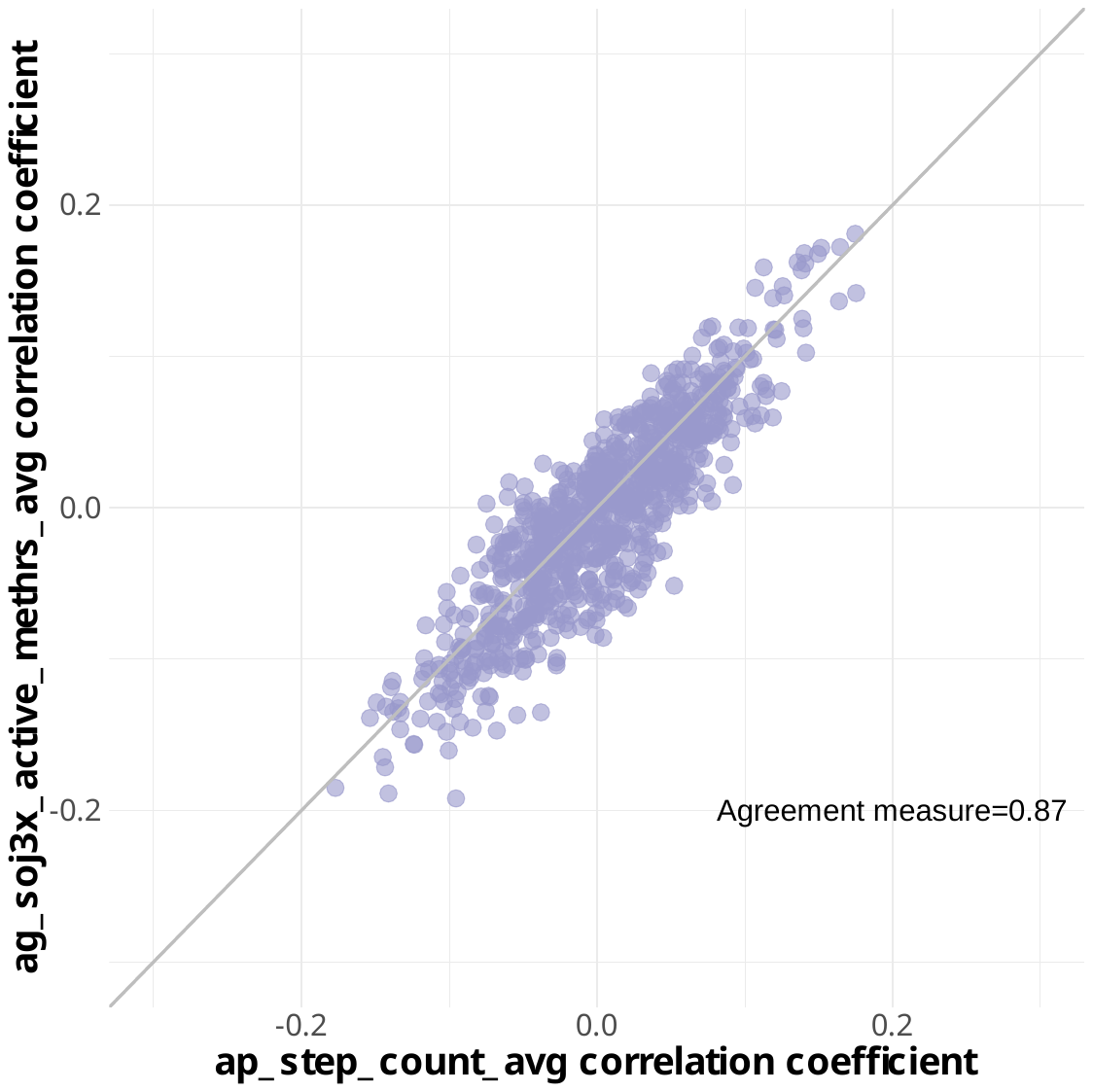

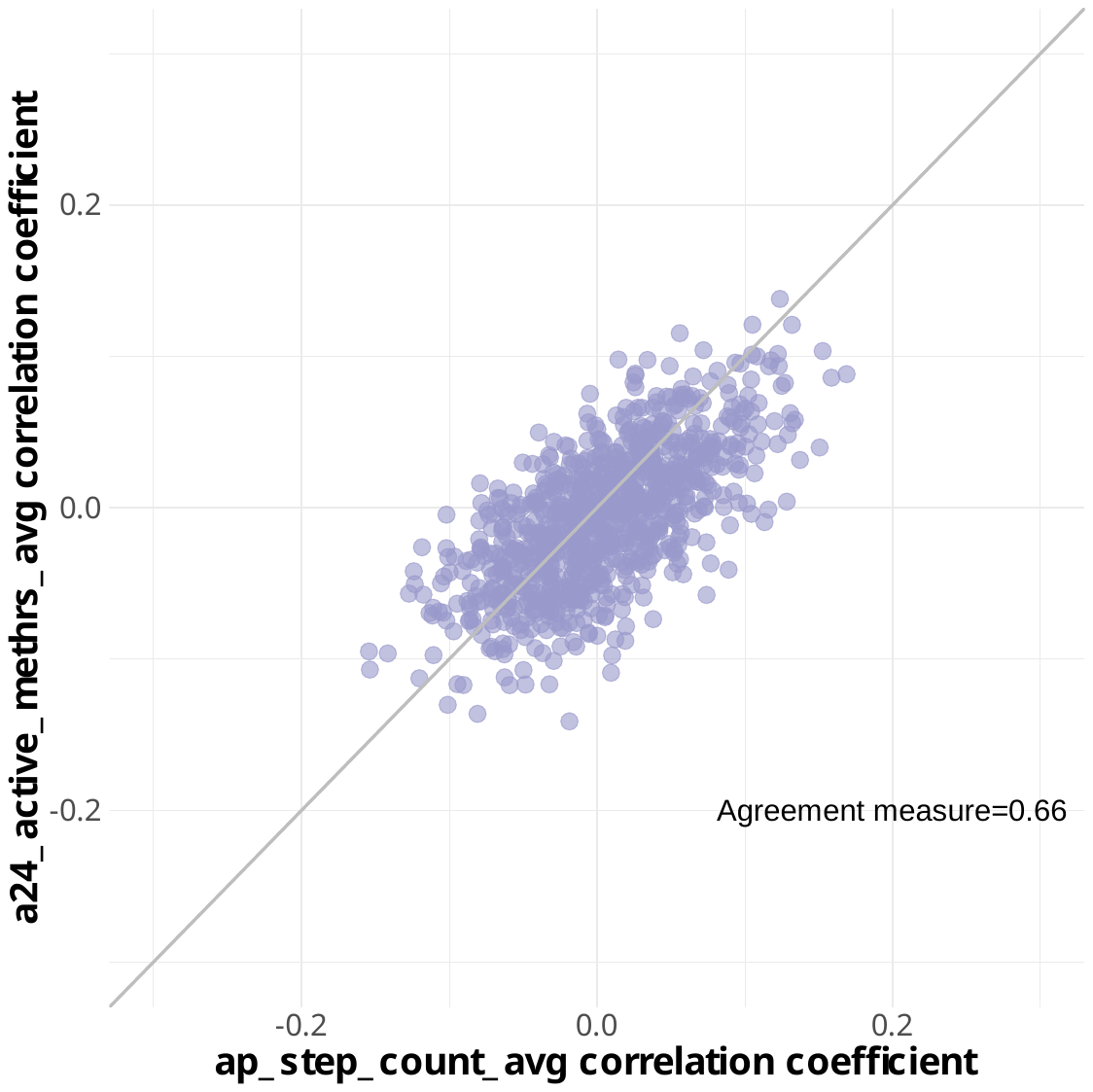

Supplementary Figure 8: Spearman partial correlations between metabolites and physical activity by assessment method, restricted to same day measures

Partial correlations adjusted for age (continuous), sex (men, women), smoking (cotinine detected: yes, no), race (non-White, White), body fat index (continuous). Agreement represents the overall correlation of each metabolite correlation with the two physical activity measures.

Abbreviations: aMET=active metabolic equivalent of task

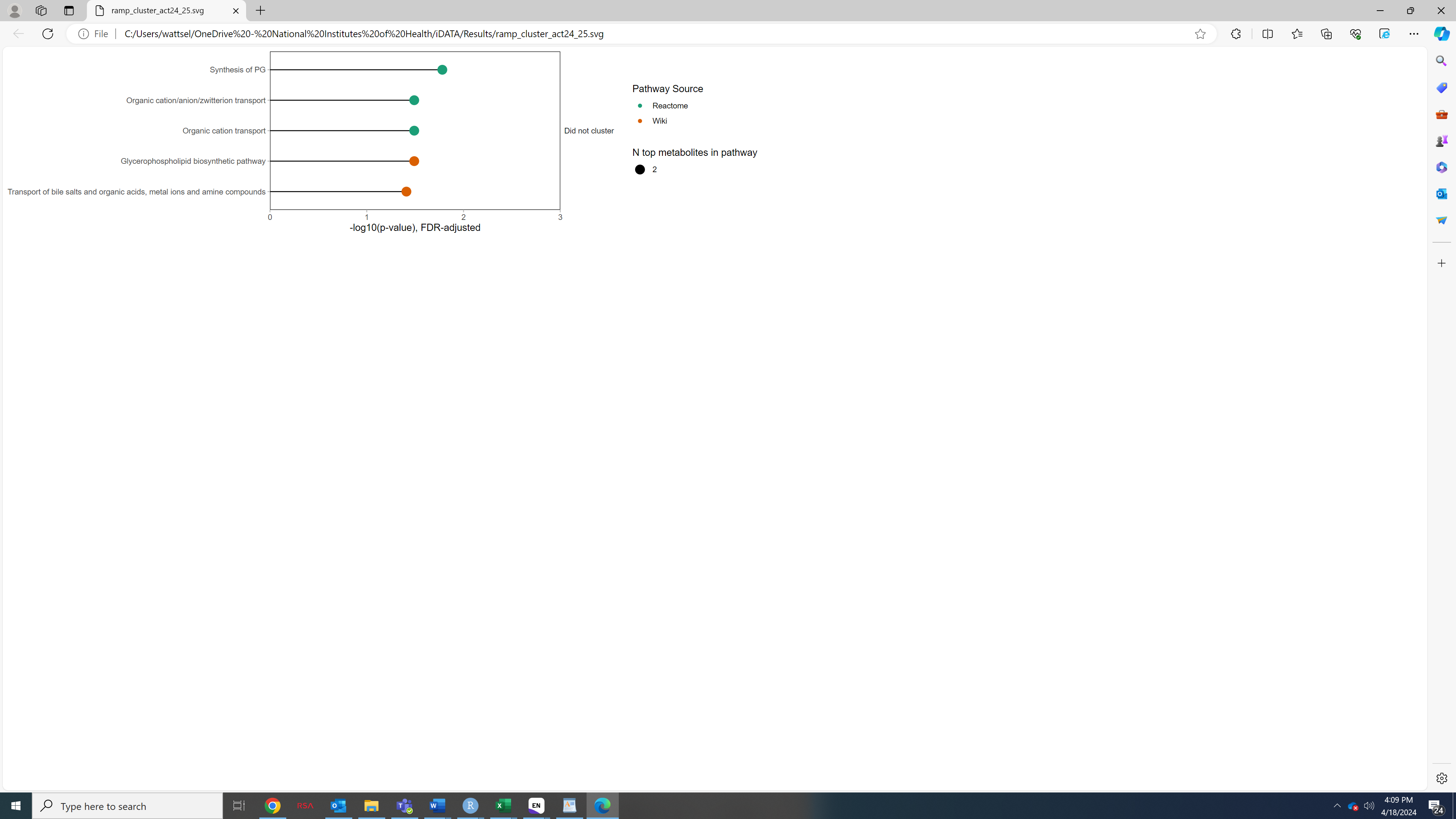

Supplementary Figure 9: Cluster pathway analysis for metabolites associated with aMET hrs/d, assessed by ACT24 using RaMP

Corrected for multiple testing using the Benjamini-Hochberg Procedure (p-adj<0.05), minimum number of pathways to cluster=3. Minimum overlap for pathways to be considered similar=20%, Minimum overlap for clusters to merge=20%. Clusters annotated to provide summary description.

Abbreviations: aMET=active metabolic equivalent of task

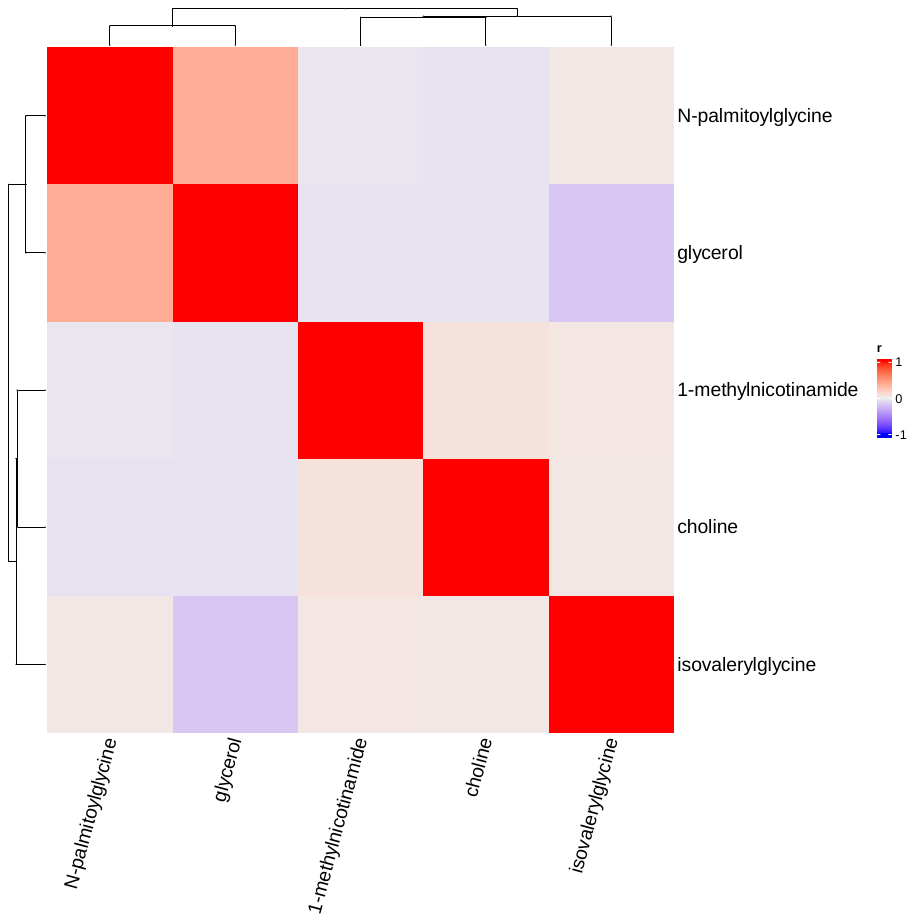

Supplementary Figure 10: Spearman correlations between serum metabolites associated with aMET hrs/day assessed by ACT24

Metabolites included were associated with aMET hrs/day, based on partial Spearman correlations, adjusted for age (continuous), sex (men, women), smoking (cotinine detected: yes, no), race (non-White, White), body fat index (continuous). Multiple testing was corrected for using the false discovery rate (p-adjusted<0.05). Heatmap created using the ComplexHeatmap R package^1^.

Abbreviations: aMET=active metabolic equivalent of task

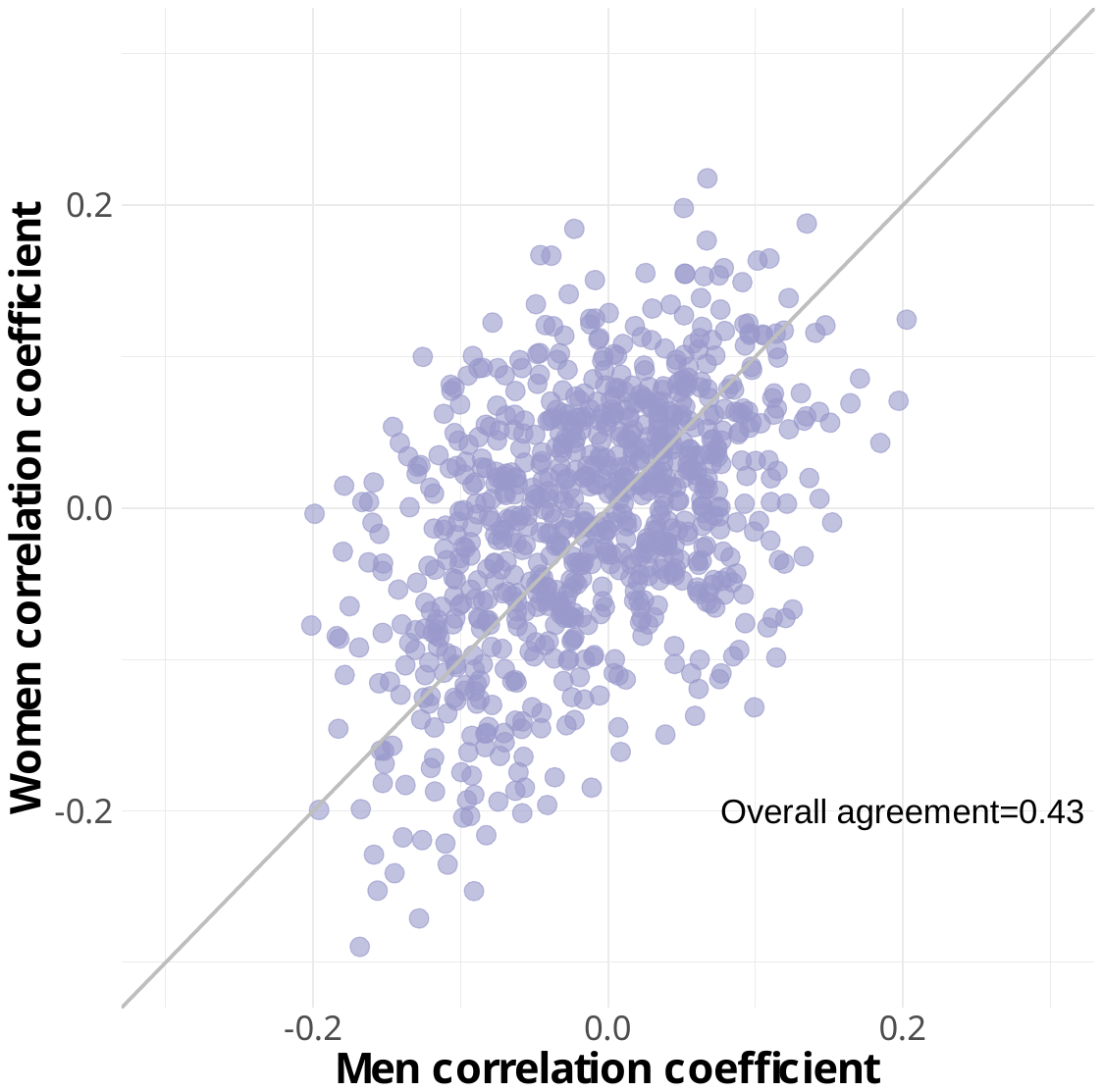

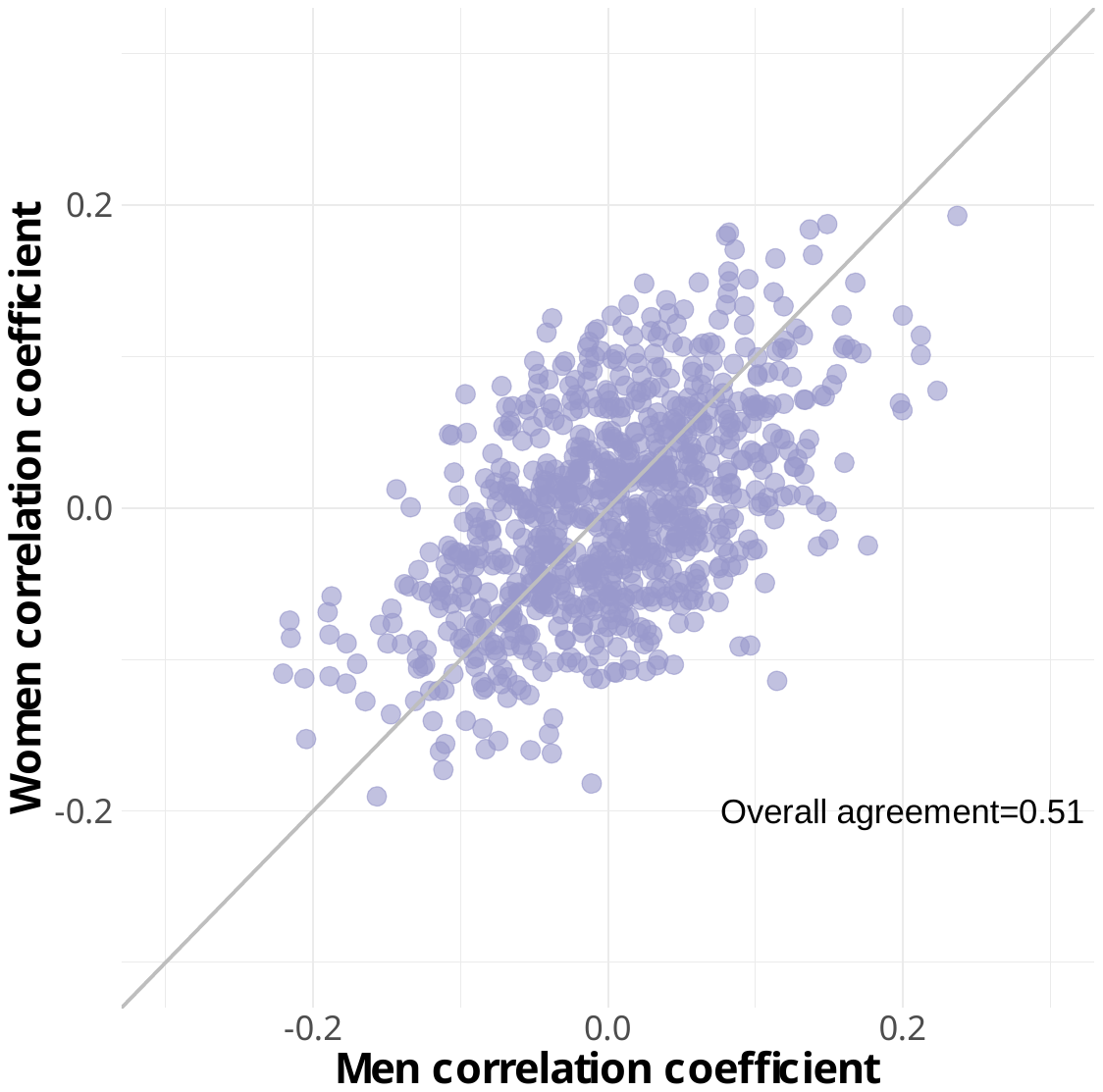

**A: Physical activity level**

**B: Step count**

**C: ActiGraph, aMET hrs/day**

**D: ACT24, aMET hrs/day**

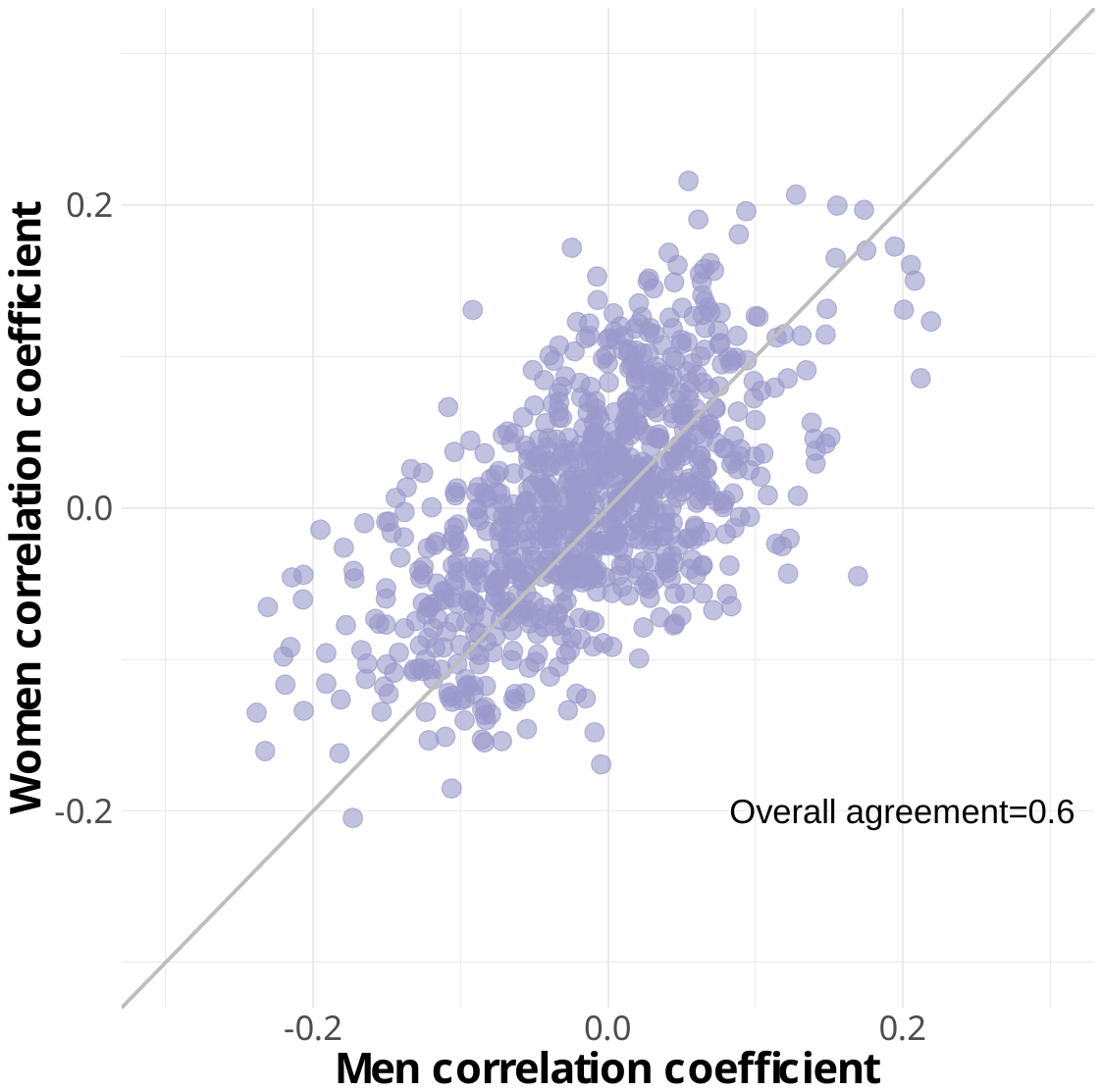

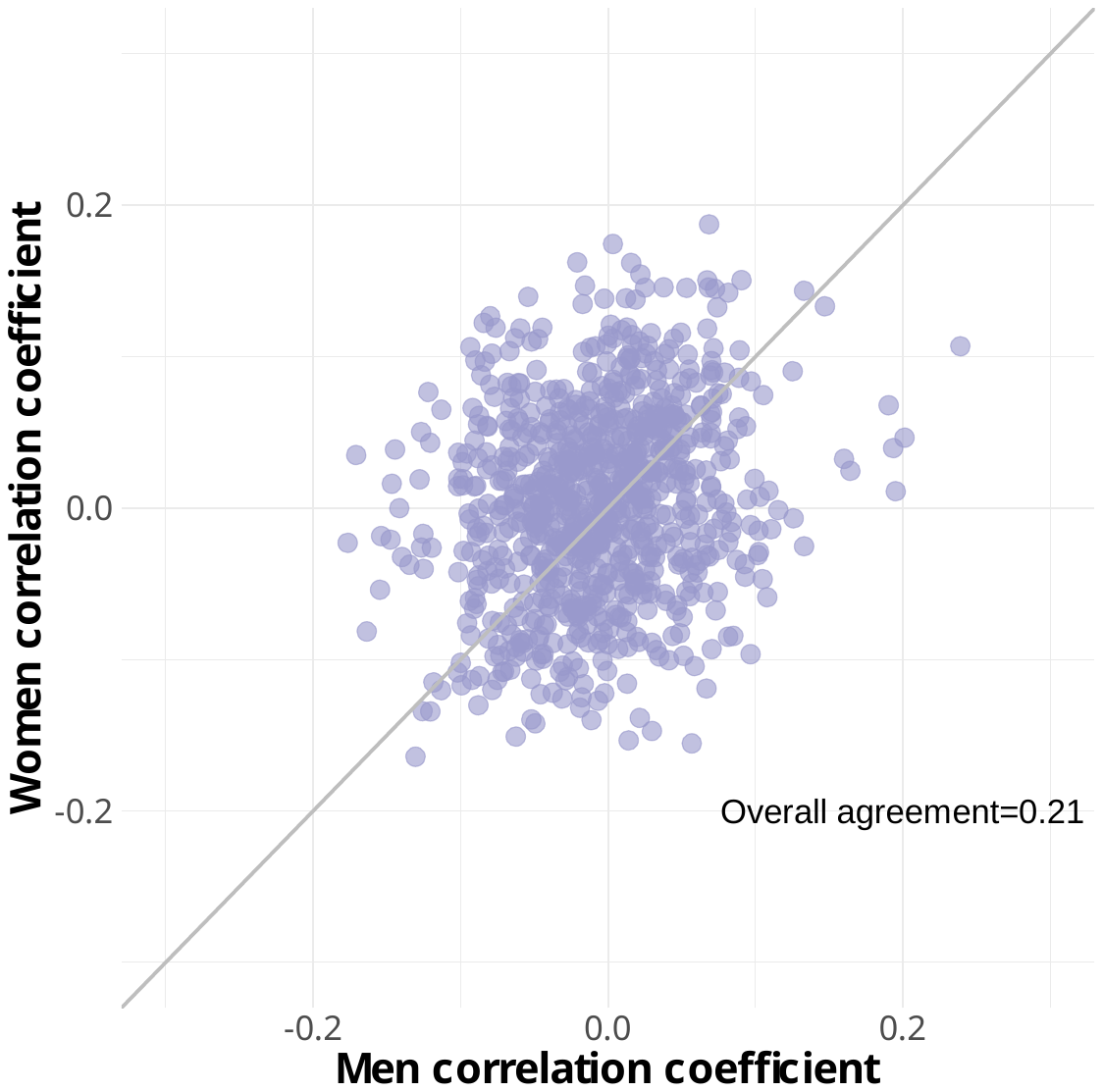

Supplementary Figure 11: Spearman partial correlations between metabolites and overall physical activity by sex

Physical activity level assessed by doubly labelled water in the IDATA study. Partial correlations adjusted for age (continuous), sex (men, women), smoking (cotinine detected: yes, no), race (non-White, White), body fat index (continuous). Overall agreement is defined as the correlation of the partial correlations.

Abbreviations: aMET= active metabolic equivalent of task, FMV=first morning void

Supplementary Table 1: IDATA study timeline of measurements

|  | **Groups 1 and 3** | | | | | | | | | | | |  | **Groups 2 and 4** | | | | | | | | | | | |
| --- | --- | --- | --- | --- | --- | --- | --- | --- | --- | --- | --- | --- | --- | --- | --- | --- | --- | --- | --- | --- | --- | --- | --- | --- | --- |
|  | **Month** | | | | | | | | | | | |  | **Month** | | | | | | | | | | | |
|  | M0 | M1 | M2 | M3 | M4 | M5 | M6 | M7 | M8 | M9 | M10 | M11 |  | M0 | M1 | M2 | M3 | M4 | M5 | M6 | M7 | M8 | M9 | M10 | M11 |
| Clinic Visit | X |  |  |  |  |  | X |  |  |  |  | X |  | X |  |  |  |  |  | X |  |  |  |  | X |
| **Biosamples** |  |  |  |  |  |  |  |  |  |  |  |  |  |  |  |  |  |  |  |  |  |  |  |  |  |
| Serum | X |  |  |  |  |  | X |  |  |  |  |  |  |  |  |  |  |  |  | X |  |  |  |  | X |
| **Physical Activity** |  |  |  |  |  |  |  |  |  |  |  |  |  |  |  |  |  |  |  |  |  |  |  |  |  |
| ActiGraph | 7d |  |  |  |  |  | 7d |  |  |  |  |  |  |  |  |  |  |  |  | 7d |  |  |  |  | 7d |
| activPAL | 7d |  |  |  |  |  | 7d |  |  |  |  |  |  |  |  |  |  |  |  | 7d |  |  |  |  | 7d |
| ACT24 | 1d |  | 1d |  | 1d |  | 1d |  | 1d |  | 1d |  |  |  | 1d |  | 1d |  |  | 1d |  | 1d |  | 1d | 1d |
| DLW | 14d |  |  |  |  |  | Sub 14d |  |  |  |  |  |  |  |  |  |  |  |  | 14d |  |  |  |  | Sub 14d |
| REE | X |  |  |  |  |  | Sub X |  |  |  |  |  |  |  |  |  |  |  |  | X |  |  |  |  | Sub X |
| **Anthropometry** |  |  |  |  |  |  |  |  |  |  |  |  |  |  |  |  |  |  |  |  |  |  |  |  |  |
| Weight, height, WC | X |  |  |  |  |  | X |  |  |  |  | X |  | X |  |  |  |  |  | X |  |  |  |  | X |
| Deuterium dilution (% body fat, FFM) | X |  |  |  |  |  | Sub X |  |  |  |  |  |  |  |  |  |  |  |  | X |  |  |  |  | Sub X |

1d = 1 previous day recall

7d = 7-day administration period

14d = 14-day administration period

Sub = repeat measurement sub study

X = clinic visit measurements

Abbreviations: DLW=doubly labelled water, FFM=fat-free mass, REE=resting energy expenditure, WC=waist circumference.

Supplementary Table 2: Characteristics of breast cancer case control study participants in the PLCO Study

|  | Cases (N=621) | |  | Controls (N=621) | |
| --- | --- | --- | --- | --- | --- |
|  | N | % |  | N | % |
| Age at blood draw, mean (SD) | 64 (5.3) |  |  | 64 (5.3) |  |
| Race |  |  |  |  |  |
| Non-Hispanic White | 555 | 90% |  | 567 | 91% |
| Other | 65 | 10% |  | 54 | 9% |
| Missing | 1 | 0.2% |  | 0 | 0% |
| Education |  |  |  |  |  |
| Up to high school | 219 | 35% |  | 213 | 34% |
| Post high school training other than college | 76 | 12% |  | 81 | 13% |
| Some college | 131 | 21% |  | 135 | 22% |
| College graduate | 109 | 18% |  | 91 | 15% |
| Postgraduate | 86 | 14% |  | 100 | 16% |
| Missing | 0 | 0% |  | 1 | 0.2% |
| Smoking |  |  |  |  |  |
| Never | 336 | 54% |  | 403 | 65% |
| Former | 237 | 38% |  | 180 | 29% |
| Current | 48 | 8% |  | 38 | 6% |
| BMI (kg/m^2^) |  |  |  |  |  |
| <25 | 202 | 33% |  | 232 | 38% |
| 25-<30 | 233 | 38% |  | 233 | 38% |
| 30+ | 178 | 29% |  | 150 | 24% |
| Missing | 8 | 1% |  | 6 | 1% |
| Healthy eating index (quartiles) | |  |  |  |  |
| 1 | 111 | 19% |  | 99 | 17% |
| 2 | 133 | 23% |  | 124 | 21% |
| 3 | 174 | 31% |  | 172 | 30% |
| 4 | 152 | 27% |  | 187 | 32% |
| Missing | 51 | 8% |  | 39 | 6% |
| Alcohol consumption (drinks/day) | |  |  |  |  |
| 0 | 107 | 19% |  | 101 | 17% |
| >0-1 | 381 | 67% |  | 401 | 69% |
| >1-2 | 48 | 8% |  | 47 | 8% |
| >2-4 | 29 | 5% |  | 28 | 5% |
| 4+ | 5 | 1% |  | 5 | 1% |
| Missing | 51 | 8% |  | 39 | 6% |
| Age at menarche |  |  |  |  |  |
| <12 | 134 | 22% |  | 112 | 18% |
| 12-13 | 323 | 52% |  | 351 | 57% |
| 14+ | 163 | 26% |  | 157 | 25% |
| Missing | 1 | 0.2% |  | 1 | 0.2% |
| Age of birth and parity |  |  |  |  |  |
| No live births | 52 | 8% |  | 57 | 9% |
| <20 years | 88 | 14% |  | 89 | 14% |
| 20-24 years, 1-2 live births | 84 | 14% |  | 62 | 10% |
| 20-24 years, >2 live births | 191 | 31% |  | 236 | 38% |
| 25-29 years, 1-2 live births | 63 | 10% |  | 51 | 8% |
| 25-29 years, >2 live births | 77 | 13% |  | 75 | 12% |
| 30+ years, 1+ live births | 60 | 10% |  | 45 | 7% |
| Missing | 6 | 1% |  | 6 | 1% |
| Menopause type |  |  |  |  |  |
| Natural menopause, <45 years | 47 | 8% |  | 37 | 6% |
| Natural menopause, 45-49 years | 106 | 17% |  | 117 | 19% |
| Natural menopause, 50-54 years | 219 | 35% |  | 220 | 36% |
| Natural menopause, 55+ years | 63 | 10% |  | 46 | 7% |
| Bilateral oophorectomy/surgery | 49 | 8% |  | 69 | 11% |
| Drug therapy/radiation | 22 | 4% |  | 10 | 2% |
| Hysterectomy, no bilateral oophorectomy | 112 | 18% |  | 120 | 19% |
| Missing | 3 | 0.4% |  | 2 | 0.3% |
| HRT use |  |  |  |  |  |
| Never | 302 | 49% |  | 300 | 48% |
| Current | 153 | 25% |  | 153 | 25% |
| Former | 166 | 27% |  | 168 | 27% |
| Benign breast disease |  |  |  |  |  |
| No | 425 | 69% |  | 471 | 77% |
| Yes | 190 | 31% |  | 142 | 23% |
| Missing | 6 | 1% |  | 8 | 1% |
| Family history of breast cancer | |  |  |  |  |
| No | 483 | 78% |  | 514 | 83% |
| Yes | 125 | 20% |  | 94 | 15% |
| Missing | 13 | 2% |  | 13 | 2% |
| Diabetes |  |  |  |  |  |
| No | 580 | 93% |  | 586 | 94% |
| Yes | 41 | 7% |  | 33 | 5% |
| Missing | 0 | 0% |  | 2 | 0.3% |

Abbreviations: BMI=body mass index, HRT=hormone replacement therapy, PLCO= Prostate, Lung, Colorectal and Ovarian Cancer Screening Trial, SD=standard deviation.

Supplementary Table 3: Physical activity associated serum metabolites by physical activity measure and super-pathway

| Super-pathway | Metabolite | PAL, DLW | |  | Steps/d, activPAL | |  | aMET hr/d, ActiGraph | |  | aMET hr/d, ACT24 | |
| --- | --- | --- | --- | --- | --- | --- | --- | --- | --- | --- | --- | --- |
|  |  | r (95% CI) | P-adj |  | r (95% CI) | P-adj |  | r (95% CI) | P-adj |  | r (95% CI) | P-adj |
| Amino Acid | N-formylmethionine | -0.14 (-0.22, -0.05) | 0.024† |  | -0.08 (-0.16, 0) | 0.19 |  | -0.14 (-0.21, -0.07) | 0.007† |  | -0.08 (-0.16, 0) | 0.3 |
|  | creatinine | -0.19 (-0.27, -0.12) | 0.001† |  | -0.1 (-0.17, -0.02) | 0.081 |  | -0.12 (-0.2, -0.04) | 0.023† |  | -0.04 (-0.12, 0.04) | 0.7 |
|  | glycine | -0.18 (-0.26, -0.1) | 0.002† |  | -0.07 (-0.15, 0) | 0.24 |  | -0.03 (-0.09, 0.05) | 0.72 |  | -0.02 (-0.1, 0.06) | 0.86 |
|  | isoleucine | -0.09 (-0.18, -0.01) | 0.14 |  | -0.11 (-0.18, -0.03) | 0.057 |  | -0.2 (-0.27, -0.13) | < 0.001† |  | -0.05 (-0.13, 0.02) | 0.61 |
|  | lysine | 0.03 (-0.04, 0.11) | 0.72 |  | 0.11 (0.03, 0.18) | 0.057 |  | 0.13 (0.06, 0.2) | 0.009† |  | 0.05 (-0.03, 0.13) | 0.61 |
|  | proline | -0.05 (-0.13, 0.04) | 0.56 |  | -0.16 (-0.23, -0.08) | 0.002† |  | -0.17 (-0.24, -0.09) | < 0.001† |  | -0.05 (-0.13, 0.02) | 0.59 |
|  | serine | -0.14 (-0.21, -0.05) | 0.028† |  | 0 (-0.07, 0.08) | 0.98 |  | -0.03 (-0.1, 0.05) | 0.67 |  | 0.01 (-0.06, 0.09) | 0.94 |
|  | glutamate | 0.06 (-0.03, 0.14) | 0.46 |  | -0.07 (-0.14, 0.01) | 0.23 |  | -0.11 (-0.18, -0.04) | 0.041† |  | -0.01 (-0.09, 0.05) | 0.93 |
|  | betaine | 0.03 (-0.06, 0.1) | 0.78 |  | 0.14 (0.07, 0.22) | 0.007† |  | 0.12 (0.05, 0.2) | 0.027† |  | 0.04 (-0.04, 0.11) | 0.73 |
|  | cysteine | -0.07 (-0.16, 0.02) | 0.34 |  | -0.15 (-0.22, -0.07) | 0.004† |  | -0.13 (-0.2, -0.05) | 0.018† |  | -0.07 (-0.15, 0.01) | 0.4 |
|  | alanine | -0.01 (-0.1, 0.07) | 0.89 |  | -0.12 (-0.19, -0.05) | 0.029† |  | -0.11 (-0.18, -0.03) | 0.049† |  | -0.03 (-0.11, 0.05) | 0.8 |
|  | sarcosine | 0.11 (0.03, 0.19) | 0.079 |  | 0.15 (0.07, 0.22) | 0.005† |  | 0.12 (0.05, 0.19) | 0.024† |  | 0.06 (-0.02, 0.13) | 0.53 |
|  | N-acetylmethionine | -0.12 (-0.2, -0.03) | 0.059 |  | -0.1 (-0.17, -0.02) | 0.1 |  | -0.12 (-0.19, -0.04) | 0.024† |  | -0.03 (-0.1, 0.05) | 0.8 |
|  | N-acetylvaline | -0.05 (-0.13, 0.03) | 0.53 |  | -0.07 (-0.15, 0.01) | 0.28 |  | -0.11 (-0.19, -0.04) | 0.033† |  | -0.04 (-0.12, 0.04) | 0.73 |
|  | N-acetylalanine | -0.12 (-0.2, -0.04) | 0.06 |  | -0.13 (-0.21, -0.05) | 0.015† |  | -0.12 (-0.2, -0.04) | 0.024† |  | -0.05 (-0.13, 0.02) | 0.57 |
|  | vanillylmandelate (VMA) | -0.12 (-0.21, -0.04) | 0.048† |  | -0.06 (-0.14, 0.01) | 0.3 |  | -0.1 (-0.17, -0.02) | 0.073 |  | -0.09 (-0.17, 0) | 0.3 |
|  | creatine | 0.12 (0.04, 0.21) | 0.048† |  | 0.08 (0.01, 0.15) | 0.17 |  | 0.09 (0.02, 0.16) | 0.098 |  | 0.08 (0, 0.16) | 0.3 |
|  | 3-methoxytyrosine | -0.12 (-0.2, -0.04) | 0.048† |  | -0.05 (-0.12, 0.03) | 0.53 |  | -0.03 (-0.11, 0.04) | 0.67 |  | -0.05 (-0.13, 0.03) | 0.62 |
|  | 3-methyl-2-oxovalerate | -0.05 (-0.14, 0.04) | 0.49 |  | -0.08 (-0.15, 0) | 0.2 |  | -0.15 (-0.23, -0.08) | 0.002† |  | -0.06 (-0.14, 0.01) | 0.49 |
|  | N-acetylaspartate (NAA) | -0.22 (-0.3, -0.14) | < 0.001† |  | -0.04 (-0.11, 0.04) | 0.58 |  | -0.09 (-0.16, -0.01) | 0.11 |  | -0.12 (-0.18, -0.04) | 0.12 |
|  | homoarginine | 0.13 (0.04, 0.21) | 0.045† |  | 0.09 (0.01, 0.16) | 0.15 |  | 0.08 (0, 0.15) | 0.15 |  | 0.06 (-0.03, 0.12) | 0.57 |
|  | N-acetylglycine | -0.14 (-0.22, -0.05) | 0.028† |  | 0.02 (-0.06, 0.1) | 0.86 |  | 0.02 (-0.06, 0.09) | 0.85 |  | 0.01 (-0.07, 0.08) | 0.97 |
|  | indolepropionate | 0.05 (-0.04, 0.13) | 0.5 |  | 0.2 (0.13, 0.27) | < 0.001† |  | 0.16 (0.08, 0.24) | < 0.001† |  | 0.04 (-0.04, 0.11) | 0.73 |
|  | 1-methyl-4-imidazoleacetate | -0.06 (-0.14, 0.02) | 0.42 |  | -0.1 (-0.17, -0.03) | 0.081 |  | -0.11 (-0.18, -0.04) | 0.033† |  | -0.05 (-0.13, 0.04) | 0.66 |
|  | N-acetylthreonine | -0.14 (-0.22, -0.06) | 0.024† |  | -0.07 (-0.15, 0) | 0.26 |  | -0.09 (-0.17, -0.02) | 0.081 |  | -0.04 (-0.12, 0.03) | 0.7 |
|  | isovalerylglycine | -0.06 (-0.15, 0.02) | 0.42 |  | 0.08 (0.01, 0.16) | 0.17 |  | 0.04 (-0.03, 0.12) | 0.5 |  | 0.14 (0.06, 0.21) | 0.039† |
|  | 2-methylbutyrylcarnitine (C5) | -0.05 (-0.14, 0.03) | 0.51 |  | -0.05 (-0.12, 0.03) | 0.52 |  | -0.12 (-0.19, -0.04) | 0.028† |  | -0.01 (-0.08, 0.07) | 0.96 |
|  | 2-hydroxy-3-methylvalerate | -0.02 (-0.11, 0.07) | 0.83 |  | -0.04 (-0.12, 0.04) | 0.57 |  | -0.13 (-0.21, -0.06) | 0.010† |  | -0.07 (-0.15, 0.01) | 0.44 |
|  | glutarylcarnitine (C5-DC) | -0.11 (-0.19, -0.03) | 0.079 |  | -0.1 (-0.17, -0.02) | 0.081 |  | -0.14 (-0.22, -0.07) | 0.007† |  | -0.07 (-0.15, 0) | 0.42 |
|  | beta-hydroxyisovaleroylcarnitine | 0.08 (0, 0.16) | 0.23 |  | 0.11 (0.04, 0.19) | 0.035† |  | 0.04 (-0.03, 0.12) | 0.49 |  | 0.03 (-0.05, 0.11) | 0.8 |
|  | tiglylcarnitine (C5:1-DC) | 0.12 (0.04, 0.21) | 0.048† |  | 0.13 (0.06, 0.21) | 0.012† |  | 0.09 (0.01, 0.16) | 0.12 |  | 0.1 (0.03, 0.17) | 0.22 |
|  | N-acetylserine | -0.12 (-0.2, -0.04) | 0.055 |  | -0.09 (-0.18, -0.02) | 0.11 |  | -0.11 (-0.19, -0.03) | 0.039† |  | -0.04 (-0.12, 0.03) | 0.71 |
|  | 4-hydroxyglutamate | 0.03 (-0.06, 0.1) | 0.78 |  | -0.12 (-0.2, -0.04) | 0.028† |  | -0.14 (-0.22, -0.07) | 0.005† |  | 0.01 (-0.06, 0.08) | 0.94 |
|  | argininate* | 0.12 (0.04, 0.2) | 0.048† |  | 0.04 (-0.03, 0.12) | 0.6 |  | 0.01 (-0.06, 0.09) | 0.87 |  | 0.07 (0, 0.14) | 0.4 |
|  | imidazole propionate | -0.08 (-0.16, 0.02) | 0.28 |  | -0.13 (-0.2, -0.05) | 0.015† |  | -0.1 (-0.17, -0.03) | 0.063 |  | -0.12 (-0.19, -0.05) | 0.091 |
|  | N-acetylcarnosine | -0.05 (-0.13, 0.03) | 0.56 |  | -0.06 (-0.14, 0.01) | 0.3 |  | -0.13 (-0.22, -0.06) | 0.010† |  | -0.02 (-0.1, 0.06) | 0.88 |
|  | N-acetyltaurine | -0.04 (-0.12, 0.05) | 0.67 |  | -0.11 (-0.18, -0.04) | 0.050† |  | -0.09 (-0.16, -0.01) | 0.12 |  | -0.02 (-0.09, 0.05) | 0.89 |
|  | p-cresol glucuronide* | -0.17 (-0.25, -0.09) | 0.004† |  | -0.05 (-0.12, 0.02) | 0.42 |  | -0.1 (-0.18, -0.03) | 0.053 |  | -0.07 (-0.14, 0) | 0.44 |
|  | C-glycosyltryptophan | -0.17 (-0.26, -0.09) | 0.004† |  | -0.12 (-0.2, -0.04) | 0.031† |  | -0.11 (-0.19, -0.03) | 0.033† |  | -0.08 (-0.16, 0) | 0.34 |
|  | (N(1) + N(8))-acetylspermidine | -0.11 (-0.19, -0.02) | 0.09 |  | -0.09 (-0.16, -0.01) | 0.13 |  | -0.11 (-0.19, -0.04) | 0.039† |  | -0.04 (-0.11, 0.04) | 0.71 |
|  | hydroxyasparagine** | -0.13 (-0.21, -0.05) | 0.029† |  | -0.13 (-0.21, -0.05) | 0.014† |  | -0.13 (-0.2, -0.05) | 0.013† |  | -0.06 (-0.14, 0.01) | 0.5 |
|  | 3-amino-2-piperidone | -0.14 (-0.22, -0.06) | 0.025† |  | -0.09 (-0.16, -0.01) | 0.14 |  | -0.08 (-0.16, 0) | 0.14 |  | -0.02 (-0.1, 0.06) | 0.88 |
|  | indoleacetoylcarnitine* | -0.01 (-0.09, 0.08) | 0.92 |  | -0.06 (-0.13, 0.01) | 0.36 |  | -0.11 (-0.18, -0.04) | 0.044† |  | 0 (-0.07, 0.08) | 0.98 |
|  | hydroxy-N6,N6,N6-trimethyllysine* | -0.14 (-0.23, -0.06) | 0.020† |  | -0.06 (-0.13, 0.01) | 0.35 |  | -0.08 (-0.15, -0.01) | 0.15 |  | -0.07 (-0.15, 0) | 0.42 |
| Carbohydrate | lactate | -0.03 (-0.12, 0.05) | 0.71 |  | -0.14 (-0.22, -0.07) | 0.005† |  | -0.2 (-0.27, -0.12) | < 0.001† |  | -0.06 (-0.13, 0.02) | 0.56 |
|  | pyruvate | 0.02 (-0.06, 0.11) | 0.83 |  | -0.13 (-0.19, -0.05) | 0.015† |  | -0.13 (-0.21, -0.06) | 0.009† |  | -0.01 (-0.09, 0.06) | 0.92 |
|  | fructose | -0.04 (-0.12, 0.04) | 0.67 |  | -0.07 (-0.14, 0.01) | 0.28 |  | -0.11 (-0.19, -0.03) | 0.049† |  | -0.01 (-0.09, 0.06) | 0.93 |
|  | N-acetylneuraminate | -0.14 (-0.22, -0.06) | 0.021† |  | -0.1 (-0.18, -0.03) | 0.062 |  | -0.1 (-0.17, -0.02) | 0.075 |  | -0.02 (-0.1, 0.05) | 0.89 |
|  | erythronate* | -0.06 (-0.16, 0.02) | 0.38 |  | -0.14 (-0.21, -0.05) | 0.009† |  | -0.12 (-0.2, -0.05) | 0.021† |  | -0.03 (-0.1, 0.05) | 0.81 |
|  | N-acetylglucosamine/N-acetylgalactosamine | -0.1 (-0.18, -0.01) | 0.11 |  | -0.11 (-0.19, -0.04) | 0.038† |  | -0.13 (-0.21, -0.06) | 0.012† |  | -0.07 (-0.14, 0.01) | 0.43 |
| Cofactors and Vitamins | 1-methylnicotinamide | 0.15 (0.06, 0.23) | 0.017† |  | 0.11 (0.03, 0.19) | 0.035† |  | 0.12 (0.04, 0.19) | 0.025† |  | 0.14 (0.07, 0.21) | 0.039† |
|  | beta-cryptoxanthin | 0.06 (-0.02, 0.14) | 0.41 |  | 0.12 (0.05, 0.19) | 0.027† |  | 0.09 (0.02, 0.17) | 0.093 |  | 0.05 (-0.02, 0.13) | 0.57 |
|  | carotene diol (1) | 0.07 (0, 0.16) | 0.33 |  | 0.13 (0.05, 0.2) | 0.014† |  | 0.14 (0.06, 0.22) | 0.007† |  | 0.07 (0, 0.15) | 0.4 |
|  | carotene diol (2) | 0.06 (-0.02, 0.15) | 0.38 |  | 0.13 (0.06, 0.2) | 0.015† |  | 0.11 (0.04, 0.19) | 0.039† |  | 0.08 (0, 0.16) | 0.35 |
| Energy | malate | -0.12 (-0.2, -0.04) | 0.049† |  | -0.11 (-0.19, -0.04) | 0.035† |  | -0.15 (-0.22, -0.08) | 0.002† |  | -0.12 (-0.19, -0.04) | 0.11 |
|  | aconitate [cis or trans] | -0.16 (-0.24, -0.08) | 0.008† |  | -0.13 (-0.21, -0.06) | 0.012† |  | -0.16 (-0.23, -0.09) | 0.001† |  | -0.06 (-0.13, 0.01) | 0.53 |
|  | 2-methylcitrate/homocitrate | 0.02 (-0.06, 0.1) | 0.81 |  | 0.12 (0.04, 0.19) | 0.025† |  | 0.08 (0, 0.16) | 0.18 |  | 0.02 (-0.06, 0.1) | 0.9 |
| Lipids | glycocholate | 0.01 (-0.09, 0.09) | 0.95 |  | -0.11 (-0.19, -0.05) | 0.036† |  | -0.14 (-0.22, -0.07) | 0.007† |  | -0.04 (-0.12, 0.03) | 0.71 |
|  | glycerol | -0.1 (-0.18, -0.01) | 0.12 |  | -0.08 (-0.15, 0.01) | 0.2 |  | -0.07 (-0.14, 0.01) | 0.23 |  | -0.14 (-0.22, -0.06) | 0.039† |
|  | choline | 0.11 (0.04, 0.19) | 0.068 |  | 0.14 (0.06, 0.21) | 0.009† |  | 0.16 (0.08, 0.22) | 0.001† |  | 0.17 (0.09, 0.24) | 0.007† |
|  | N-palmitoyl-sphingosine (d18:1/16:0) | -0.11 (-0.19, -0.02) | 0.079 |  | -0.13 (-0.2, -0.05) | 0.015† |  | -0.08 (-0.16, -0.01) | 0.13 |  | -0.02 (-0.09, 0.06) | 0.91 |
|  | stearoyl sphingomyelin (d18:1/18:0) | -0.14 (-0.22, -0.06) | 0.022† |  | -0.04 (-0.11, 0.03) | 0.58 |  | -0.06 (-0.12, 0.01) | 0.33 |  | -0.06 (-0.14, 0.01) | 0.49 |
|  | 1-palmityl-GPC (O-16:0) | -0.12 (-0.21, -0.04) | 0.048† |  | 0.06 (-0.02, 0.13) | 0.4 |  | 0.03 (-0.04, 0.1) | 0.71 |  | 0.03 (-0.05, 0.1) | 0.81 |
|  | sphingosine 1-phosphate | -0.04 (-0.12, 0.04) | 0.6 |  | -0.06 (-0.13, 0.02) | 0.37 |  | -0.12 (-0.19, -0.04) | 0.027† |  | 0.02 (-0.05, 0.1) | 0.86 |
|  | epiandrosterone sulfate | 0.1 (0.02, 0.17) | 0.13 |  | 0.13 (0.05, 0.2) | 0.015† |  | 0.08 (0, 0.16) | 0.14 |  | 0.06 (-0.01, 0.13) | 0.52 |
|  | 1-dihomo-linolenoyl-GPC (20:3n3 or 6)* | 0.14 (0.06, 0.22) | 0.024† |  | 0.07 (-0.01, 0.14) | 0.25 |  | 0.07 (0, 0.14) | 0.23 |  | 0.06 (-0.01, 0.14) | 0.49 |
|  | cholesterol sulfate | -0.07 (-0.15, 0.02) | 0.34 |  | -0.11 (-0.18, -0.04) | 0.037† |  | -0.06 (-0.13, 0.01) | 0.28 |  | 0.01 (-0.06, 0.09) | 0.97 |
|  | 7alpha-hydroxy-3-oxo-4-cholestenoate (7-Hoca) | -0.07 (-0.15, 0.01) | 0.35 |  | -0.07 (-0.15, 0) | 0.23 |  | -0.11 (-0.19, -0.04) | 0.046† |  | -0.12 (-0.19, -0.05) | 0.091 |
|  | hexadecanedioate (C16-DC) | -0.13 (-0.21, -0.05) | 0.034† |  | -0.08 (-0.14, -0.01) | 0.21 |  | -0.1 (-0.18, -0.03) | 0.061 |  | -0.08 (-0.15, -0.01) | 0.3 |
|  | hexanoylglutamine | -0.12 (-0.21, -0.04) | 0.049† |  | -0.03 (-0.11, 0.05) | 0.68 |  | -0.07 (-0.15, 0.01) | 0.23 |  | -0.03 (-0.1, 0.04) | 0.81 |
|  | sphinganine-1-phosphate | -0.06 (-0.13, 0.02) | 0.45 |  | -0.04 (-0.11, 0.03) | 0.58 |  | -0.12 (-0.18, -0.04) | 0.028† |  | 0 (-0.07, 0.07) | 0.98 |
|  | sphingomyelin (d18:1/18:1, d18:2/18:0) | -0.17 (-0.25, -0.09) | 0.004† |  | -0.07 (-0.14, 0) | 0.24 |  | -0.05 (-0.12, 0.02) | 0.43 |  | -0.07 (-0.15, 0.01) | 0.43 |
|  | palmitoyl sphingomyelin (d18:1/16:0) | -0.15 (-0.23, -0.07) | 0.012† |  | -0.04 (-0.11, 0.04) | 0.58 |  | -0.05 (-0.12, 0.02) | 0.38 |  | -0.03 (-0.11, 0.05) | 0.8 |
|  | 16alpha-hydroxy DHEA 3-sulfate | -0.08 (-0.17, 0) | 0.22 |  | -0.11 (-0.19, -0.04) | 0.036† |  | -0.17 (-0.24, -0.09) | < 0.001† |  | -0.07 (-0.14, 0.01) | 0.48 |
|  | andro steroid monosulfate C19H28O6S (1)* | -0.05 (-0.14, 0.03) | 0.49 |  | -0.09 (-0.16, 0) | 0.15 |  | -0.15 (-0.22, -0.07) | 0.004† |  | -0.06 (-0.13, 0.01) | 0.54 |
|  | N-palmitoylglycine | -0.12 (-0.2, -0.03) | 0.064 |  | -0.1 (-0.17, -0.03) | 0.081 |  | -0.07 (-0.14, 0.01) | 0.27 |  | -0.15 (-0.22, -0.08) | 0.022† |
|  | sphingomyelin (d18:2/16:0, d18:1/16:1)* | -0.17 (-0.26, -0.09) | 0.004† |  | -0.03 (-0.1, 0.06) | 0.74 |  | -0.02 (-0.09, 0.05) | 0.81 |  | -0.03 (-0.11, 0.04) | 0.78 |
|  | 2-aminoheptanoate | -0.04 (-0.13, 0.04) | 0.58 |  | -0.12 (-0.2, -0.04) | 0.028† |  | -0.1 (-0.17, -0.02) | 0.073 |  | -0.08 (-0.16, -0.01) | 0.3 |
|  | 1-dihomo-linolenoyl-GPE (20:3n3 or 6)* | 0.16 (0.08, 0.24) | 0.007† |  | 0.02 (-0.06, 0.09) | 0.81 |  | 0.06 (-0.02, 0.13) | 0.34 |  | 0.05 (-0.03, 0.13) | 0.62 |
|  | sphingomyelin (d18:1/24:1, d18:2/24:0)* | -0.13 (-0.22, -0.05) | 0.030† |  | -0.01 (-0.08, 0.07) | 0.96 |  | 0.03 (-0.05, 0.1) | 0.72 |  | -0.01 (-0.08, 0.07) | 0.98 |
|  | sphingomyelin (d18:1/20:1, d18:2/20:0)* | -0.15 (-0.22, -0.06) | 0.015† |  | 0.01 (-0.06, 0.09) | 0.88 |  | 0.03 (-0.05, 0.1) | 0.74 |  | 0 (-0.08, 0.08) | 0.98 |
|  | sphingomyelin (d18:1/20:2, d18:2/20:1, d16:1/22:2)* | -0.14 (-0.22, -0.05) | 0.022† |  | -0.04 (-0.11, 0.04) | 0.58 |  | -0.02 (-0.1, 0.05) | 0.76 |  | -0.04 (-0.13, 0.03) | 0.7 |
|  | sphingomyelin (d18:1/22:2, d18:2/22:1, d16:1/24:2)* | -0.17 (-0.25, -0.08) | 0.005† |  | -0.02 (-0.09, 0.06) | 0.82 |  | -0.01 (-0.09, 0.07) | 0.92 |  | -0.03 (-0.11, 0.05) | 0.81 |
|  | adipoylcarnitine (C6-DC) | -0.13 (-0.21, -0.04) | 0.042† |  | -0.05 (-0.13, 0.03) | 0.44 |  | -0.06 (-0.13, 0.01) | 0.32 |  | 0 (-0.08, 0.08) | 0.98 |
|  | sphingomyelin (d18:1/17:0, d17:1/18:0, d19:1/16:0) | -0.16 (-0.24, -0.09) | 0.005† |  | -0.02 (-0.09, 0.05) | 0.8 |  | -0.03 (-0.11, 0.04) | 0.63 |  | -0.04 (-0.12, 0.03) | 0.72 |
|  | sphingomyelin (d18:2/24:1, d18:1/24:2)* | -0.19 (-0.28, -0.1) | 0.001† |  | -0.04 (-0.11, 0.03) | 0.55 |  | -0.01 (-0.08, 0.06) | 0.91 |  | -0.05 (-0.13, 0.03) | 0.65 |
|  | 1-palmitoyl-2-dihomo-linolenoyl-GPC (16:0/20:3n3 or 6)* | 0.12 (0.04, 0.21) | 0.049† |  | 0.01 (-0.06, 0.08) | 0.9 |  | 0.03 (-0.04, 0.11) | 0.63 |  | 0.05 (-0.03, 0.11) | 0.68 |
|  | 1-(1-enyl-stearoyl)-2-docosahexaenoyl-GPE (P-18:0/22:6)* | -0.13 (-0.21, -0.04) | 0.039† |  | -0.01 (-0.09, 0.07) | 0.94 |  | -0.02 (-0.09, 0.06) | 0.81 |  | -0.01 (-0.09, 0.06) | 0.95 |
|  | 1-(1-enyl-stearoyl)-2-docosahexaenoyl-GPC (P-18:0/22:6)* | -0.14 (-0.21, -0.05) | 0.028† |  | 0.01 (-0.08, 0.08) | 0.96 |  | -0.02 (-0.09, 0.06) | 0.82 |  | 0 (-0.08, 0.07) | 0.98 |
|  | 1-stearyl-2-arachidonoyl-GPC (O-18:0/20:4)* | -0.14 (-0.23, -0.06) | 0.024† |  | 0 (-0.08, 0.07) | 0.96 |  | -0.04 (-0.11, 0.03) | 0.56 |  | -0.02 (-0.1, 0.05) | 0.85 |
|  | sphingomyelin (d18:1/21:0, d17:1/22:0, d16:1/23:0)* | 0.04 (-0.04, 0.13) | 0.58 |  | 0.12 (0.05, 0.19) | 0.021† |  | 0.14 (0.06, 0.21) | 0.007† |  | 0.11 (0.04, 0.19) | 0.13 |
|  | sphingomyelin (d18:0/18:0, d19:0/17:0)* | -0.07 (-0.16, 0.02) | 0.35 |  | -0.11 (-0.19, -0.04) | 0.036† |  | -0.11 (-0.18, -0.03) | 0.044† |  | -0.06 (-0.13, 0.02) | 0.56 |
|  | lactosyl-N-palmitoyl-sphingosine (d18:1/16:0) | -0.13 (-0.21, -0.05) | 0.032† |  | -0.09 (-0.16, -0.01) | 0.14 |  | -0.11 (-0.18, -0.03) | 0.043† |  | -0.04 (-0.12, 0.04) | 0.74 |
|  | 1-stearoyl-2-dihomo-linolenoyl-GPC (18:0/20:3n3 or 6)* | 0.13 (0.04, 0.21) | 0.044† |  | -0.02 (-0.1, 0.05) | 0.74 |  | -0.01 (-0.09, 0.06) | 0.87 |  | 0.03 (-0.05, 0.1) | 0.81 |
|  | 1-linoleoyl-2-arachidonoyl-GPC (18:2/20:4n6)* | 0.07 (-0.01, 0.15) | 0.33 |  | 0.16 (0.09, 0.23) | 0.002† |  | 0.18 (0.1, 0.25) | < 0.001† |  | 0.07 (0, 0.15) | 0.42 |
|  | 1-myristoyl-2-arachidonoyl-GPC (14:0/20:4)* | 0.1 (0.02, 0.19) | 0.1 |  | 0.08 (0.01, 0.15) | 0.19 |  | 0.12 (0.04, 0.19) | 0.028† |  | 0.03 (-0.04, 0.1) | 0.8 |
|  | 1-stearyl-GPC (O-18:0)* | -0.14 (-0.23, -0.06) | 0.024† |  | 0.01 (-0.06, 0.09) | 0.88 |  | 0 (-0.07, 0.07) | 1 |  | -0.03 (-0.11, 0.05) | 0.78 |
|  | 1-stearoyl-2-dihomo-linolenoyl-GPE (18:0/20:3n3 or 6)* | 0.13 (0.04, 0.21) | 0.039† |  | -0.02 (-0.1, 0.05) | 0.77 |  | -0.03 (-0.1, 0.05) | 0.72 |  | 0.03 (-0.05, 0.1) | 0.79 |
|  | palmitoylcholine | 0.02 (-0.07, 0.11) | 0.84 |  | 0.15 (0.08, 0.23) | 0.003† |  | 0.17 (0.1, 0.24) | < 0.001† |  | 0.09 (0.02, 0.18) | 0.24 |
|  | glycosyl-N-palmitoyl-sphingosine (d18:1/16:0) | -0.15 (-0.23, -0.06) | 0.013† |  | -0.09 (-0.16, -0.02) | 0.12 |  | -0.07 (-0.14, 0) | 0.23 |  | -0.04 (-0.12, 0.03) | 0.71 |
|  | oleoylcholine | 0.04 (-0.04, 0.12) | 0.6 |  | 0.14 (0.06, 0.2) | 0.010† |  | 0.16 (0.09, 0.24) | < 0.001† |  | 0.1 (0.02, 0.18) | 0.23 |
|  | arachidonoylcholine | 0.01 (-0.08, 0.09) | 0.93 |  | 0.16 (0.09, 0.24) | 0.002† |  | 0.18 (0.11, 0.26) | < 0.001† |  | 0.06 (-0.02, 0.14) | 0.52 |
|  | docosahexaenoylcholine | -0.01 (-0.1, 0.08) | 0.92 |  | 0.11 (0.04, 0.19) | 0.036† |  | 0.11 (0.04, 0.19) | 0.033† |  | 0.05 (-0.03, 0.13) | 0.61 |
|  | palmitoloelycholine | 0.08 (0, 0.17) | 0.22 |  | 0.09 (0.01, 0.16) | 0.12 |  | 0.13 (0.05, 0.2) | 0.013† |  | 0.06 (-0.01, 0.13) | 0.56 |
|  | dihomo-linolenoyl-choline | 0.12 (0.03, 0.2) | 0.055 |  | 0.14 (0.06, 0.21) | 0.009† |  | 0.15 (0.07, 0.22) | 0.003† |  | 0.1 (0.03, 0.18) | 0.19 |
|  | eicosapentaenoylcholine | 0.06 (-0.02, 0.15) | 0.38 |  | 0.15 (0.08, 0.23) | 0.004† |  | 0.18 (0.1, 0.25) | < 0.001† |  | 0.06 (-0.01, 0.14) | 0.54 |
|  | 1-palmityl-2-palmitoyl-GPC (O-16:0/16:0)* | -0.12 (-0.21, -0.04) | 0.048† |  | -0.05 (-0.12, 0.02) | 0.46 |  | -0.03 (-0.1, 0.05) | 0.69 |  | -0.04 (-0.12, 0.03) | 0.69 |
|  | glycerophosphoserine* | -0.04 (-0.13, 0.05) | 0.61 |  | -0.09 (-0.16, -0.02) | 0.11 |  | -0.11 (-0.19, -0.04) | 0.031† |  | -0.02 (-0.09, 0.06) | 0.91 |
|  | stearoylcholine* | 0.04 (-0.05, 0.13) | 0.64 |  | 0.16 (0.1, 0.23) | 0.002† |  | 0.18 (0.11, 0.25) | < 0.001† |  | 0.12 (0.05, 0.2) | 0.091 |
|  | linoleoylcholine* | 0.02 (-0.07, 0.1) | 0.88 |  | 0.16 (0.09, 0.23) | 0.003† |  | 0.18 (0.11, 0.24) | < 0.001† |  | 0.13 (0.06, 0.2) | 0.091 |
|  | sphingomyelin (d18:2/18:1)* | -0.17 (-0.25, -0.1) | 0.004† |  | -0.05 (-0.12, 0.03) | 0.48 |  | -0.01 (-0.08, 0.06) | 0.92 |  | -0.03 (-0.11, 0.05) | 0.8 |
|  | sphingomyelin (d18:2/24:2)* | -0.22 (-0.3, -0.13) | < 0.001† |  | -0.06 (-0.13, 0.01) | 0.31 |  | -0.03 (-0.11, 0.04) | 0.63 |  | -0.06 (-0.13, 0.02) | 0.56 |
|  | sphingomyelin (d18:2/23:1)* | -0.18 (-0.27, -0.1) | 0.002† |  | -0.03 (-0.1, 0.05) | 0.72 |  | -0.01 (-0.09, 0.06) | 0.87 |  | -0.02 (-0.11, 0.05) | 0.88 |
|  | lignoceroylcarnitine (C24)* | 0.14 (0.05, 0.22) | 0.028† |  | 0.08 (0.01, 0.16) | 0.17 |  | 0.12 (0.04, 0.19) | 0.028† |  | 0.12 (0.05, 0.2) | 0.091 |
|  | octadecenedioate (C18:1-DC) | -0.14 (-0.22, -0.06) | 0.028† |  | -0.05 (-0.13, 0.02) | 0.43 |  | -0.07 (-0.14, 0) | 0.23 |  | -0.05 (-0.12, 0.02) | 0.6 |
|  | octadecadienedioate (C18:2-DC)* | -0.12 (-0.2, -0.05) | 0.047† |  | -0.04 (-0.11, 0.04) | 0.58 |  | -0.03 (-0.1, 0.05) | 0.71 |  | 0 (-0.07, 0.08) | 0.99 |
|  | palmitoyl-sphingosine-phosphoethanolamine (d18:1/16:0) | -0.14 (-0.22, -0.06) | 0.023† |  | -0.05 (-0.12, 0.03) | 0.51 |  | -0.03 (-0.1, 0.04) | 0.69 |  | -0.04 (-0.12, 0.03) | 0.7 |
|  | branched chain 14:0 dicarboxylic acid** | 0.06 (-0.02, 0.14) | 0.4 |  | 0.12 (0.05, 0.19) | 0.025† |  | 0.07 (0, 0.15) | 0.22 |  | -0.01 (-0.08, 0.07) | 0.95 |
| Nucleotide | uridine | 0.13 (0.04, 0.21) | 0.039† |  | 0.09 (0.02, 0.16) | 0.13 |  | 0.06 (-0.03, 0.13) | 0.36 |  | 0.03 (-0.04, 0.11) | 0.77 |
|  | pseudouridine | -0.16 (-0.24, -0.08) | 0.010† |  | -0.04 (-0.12, 0.03) | 0.58 |  | -0.07 (-0.15, 0) | 0.23 |  | -0.07 (-0.15, 0.01) | 0.44 |
|  | Uracil | 0.13 (0.05, 0.21) | 0.037† |  | 0.09 (0.01, 0.16) | 0.15 |  | 0.09 (0.02, 0.17) | 0.089 |  | 0 (-0.08, 0.08) | 0.98 |
|  | N1-methyladenosine | -0.09 (-0.16, 0) | 0.2 |  | -0.14 (-0.22, -0.07) | 0.006† |  | -0.14 (-0.22, -0.06) | 0.007† |  | -0.06 (-0.14, 0.01) | 0.49 |
|  | N4-acetylcytidine | -0.07 (-0.15, 0.03) | 0.35 |  | -0.12 (-0.19, -0.04) | 0.028† |  | -0.15 (-0.22, -0.08) | 0.002† |  | -0.11 (-0.18, -0.03) | 0.16 |
|  | N6-carbamoylthreonyladenosine | -0.13 (-0.22, -0.06) | 0.029† |  | -0.08 (-0.16, -0.01) | 0.17 |  | -0.12 (-0.19, -0.04) | 0.028† |  | -0.06 (-0.14, 0.02) | 0.53 |
|  | orotidine | -0.11 (-0.19, -0.01) | 0.099 |  | -0.11 (-0.19, -0.03) | 0.040† |  | -0.11 (-0.18, -0.03) | 0.041† |  | -0.04 (-0.11, 0.03) | 0.75 |
|  | 5,6-dihydrouridine | -0.14 (-0.23, -0.06) | 0.022† |  | -0.09 (-0.16, -0.01) | 0.12 |  | -0.12 (-0.2, -0.04) | 0.025† |  | -0.1 (-0.17, -0.02) | 0.23 |
|  | 3-(3-amino-3-carboxypropyl)uridine* | -0.12 (-0.2, -0.04) | 0.055 |  | -0.11 (-0.2, -0.04) | 0.040† |  | -0.12 (-0.19, -0.04) | 0.028† |  | -0.08 (-0.15, 0) | 0.38 |
| Peptide | gamma-glutamylvaline | -0.06 (-0.15, 0.02) | 0.42 |  | -0.11 (-0.18, -0.03) | 0.043† |  | -0.11 (-0.19, -0.05) | 0.033† |  | -0.04 (-0.11, 0.04) | 0.73 |
|  | pyroglutamylglycine | -0.01 (-0.09, 0.08) | 0.91 |  | -0.01 (-0.09, 0.06) | 0.88 |  | -0.11 (-0.19, -0.03) | 0.048† |  | -0.01 (-0.09, 0.06) | 0.94 |
|  | gamma-glutamylglycine | -0.18 (-0.27, -0.1) | 0.002† |  | -0.08 (-0.16, 0) | 0.19 |  | -0.03 (-0.11, 0.05) | 0.63 |  | -0.03 (-0.1, 0.05) | 0.81 |
|  | gamma-glutamylthreonine | -0.13 (-0.21, -0.04) | 0.042† |  | -0.04 (-0.12, 0.03) | 0.57 |  | -0.01 (-0.08, 0.07) | 0.94 |  | -0.02 (-0.09, 0.06) | 0.89 |
|  | phenylacetylglutamine | -0.13 (-0.22, -0.04) | 0.029† |  | -0.04 (-0.11, 0.04) | 0.63 |  | -0.1 (-0.16, -0.02) | 0.079 |  | -0.04 (-0.11, 0.03) | 0.73 |
|  | gamma-glutamylisoleucine* | -0.08 (-0.16, 0) | 0.26 |  | -0.12 (-0.19, -0.04) | 0.033† |  | -0.17 (-0.25, -0.1) | < 0.001† |  | -0.02 (-0.1, 0.05) | 0.88 |
|  | prolylproline | -0.09 (-0.17, 0) | 0.19 |  | -0.13 (-0.21, -0.06) | 0.012† |  | -0.12 (-0.19, -0.05) | 0.024† |  | -0.03 (-0.11, 0.04) | 0.77 |
|  | 4-hydroxyphenylacetylglutamine | -0.09 (-0.17, 0) | 0.19 |  | -0.11 (-0.19, -0.04) | 0.038† |  | -0.11 (-0.18, -0.03) | 0.044† |  | -0.09 (-0.17, -0.01) | 0.3 |
|  | gamma-glutamylcitrulline* | -0.01 (-0.1, 0.07) | 0.89 |  | 0.12 (0.05, 0.19) | 0.026† |  | 0.11 (0.03, 0.19) | 0.047† |  | 0.09 (0.02, 0.18) | 0.29 |
| Unknown/PC | glutamine_degradant* | -0.15 (-0.23, -0.06) | 0.014† |  | -0.09 (-0.16, -0.02) | 0.14 |  | -0.1 (-0.18, -0.03) | 0.055 |  | -0.08 (-0.16, -0.01) | 0.3 |
|  | glutamine conjugate of C6H10O2 (2)* | -0.13 (-0.21, -0.05) | 0.041† |  | -0.08 (-0.15, 0) | 0.16 |  | -0.1 (-0.17, -0.02) | 0.067 |  | -0.08 (-0.15, -0.01) | 0.3 |
|  | X-11315 | 0.11 (0.03, 0.19) | 0.091 |  | 0.13 (0.06, 0.2) | 0.015† |  | 0.12 (0.04, 0.19) | 0.028† |  | 0.05 (-0.03, 0.13) | 0.68 |
|  | X-11852 | 0.07 (-0.02, 0.14) | 0.37 |  | 0.11 (0.04, 0.18) | 0.040† |  | 0.09 (0.01, 0.16) | 0.11 |  | 0.05 (-0.02, 0.13) | 0.59 |
|  | X-12026 | -0.16 (-0.25, -0.08) | 0.006† |  | -0.07 (-0.15, 0.01) | 0.29 |  | -0.08 (-0.15, 0) | 0.16 |  | -0.03 (-0.11, 0.04) | 0.77 |
|  | X-12117 | -0.15 (-0.23, -0.07) | 0.013† |  | -0.18 (-0.26, -0.11) | < 0.001† |  | -0.19 (-0.27, -0.12) | < 0.001† |  | -0.04 (-0.12, 0.04) | 0.76 |
|  | X-12283 | 0.06 (-0.02, 0.14) | 0.45 |  | 0.13 (0.06, 0.2) | 0.015† |  | 0.1 (0.02, 0.17) | 0.079 |  | 0.01 (-0.07, 0.07) | 0.98 |
|  | X-12306 | 0.17 (0.08, 0.25) | 0.004† |  | 0.11 (0.04, 0.18) | 0.036† |  | 0.1 (0.02, 0.18) | 0.067 |  | 0.04 (-0.03, 0.12) | 0.69 |
|  | X-12544 | 0.03 (-0.06, 0.11) | 0.76 |  | 0.13 (0.05, 0.2) | 0.015† |  | 0.03 (-0.03, 0.11) | 0.62 |  | 0.03 (-0.04, 0.11) | 0.77 |
|  | X-14056 | -0.02 (-0.11, 0.06) | 0.83 |  | -0.16 (-0.23, -0.08) | 0.002† |  | -0.13 (-0.2, -0.05) | 0.013† |  | -0.06 (-0.13, 0.01) | 0.53 |
|  | X-15245 | -0.03 (-0.11, 0.05) | 0.78 |  | -0.15 (-0.21, -0.07) | 0.005† |  | -0.17 (-0.24, -0.09) | < 0.001† |  | -0.06 (-0.14, 0.01) | 0.49 |
|  | X-21258 | 0.06 (-0.03, 0.15) | 0.43 |  | 0.16 (0.08, 0.23) | 0.002† |  | 0.13 (0.05, 0.21) | 0.010† |  | 0.08 (0.01, 0.16) | 0.3 |
|  | X-21310 | -0.12 (-0.21, -0.04) | 0.049† |  | -0.03 (-0.11, 0.04) | 0.69 |  | -0.1 (-0.18, -0.02) | 0.067 |  | -0.04 (-0.13, 0.04) | 0.71 |
|  | X-22162 | 0.08 (0, 0.16) | 0.24 |  | 0.09 (0.02, 0.16) | 0.11 |  | 0.11 (0.03, 0.19) | 0.033† |  | 0.03 (-0.04, 0.1) | 0.79 |
|  | X-23666 | -0.17 (-0.25, -0.09) | 0.005† |  | -0.08 (-0.15, -0.01) | 0.19 |  | -0.1 (-0.18, -0.03) | 0.055 |  | -0.05 (-0.12, 0.03) | 0.67 |
|  | X-24337 | -0.02 (-0.1, 0.07) | 0.87 |  | -0.16 (-0.24, -0.09) | 0.002† |  | -0.12 (-0.2, -0.04) | 0.024† |  | -0.04 (-0.12, 0.04) | 0.74 |
|  | X-25343 | 0.01 (-0.08, 0.09) | 0.92 |  | -0.15 (-0.22, -0.07) | 0.004† |  | -0.13 (-0.2, -0.06) | 0.013† |  | -0.03 (-0.11, 0.04) | 0.79 |
|  | X-25520 | 0.05 (-0.03, 0.13) | 0.5 |  | 0.11 (0.04, 0.19) | 0.035† |  | 0.06 (-0.02, 0.14) | 0.34 |  | -0.02 (-0.1, 0.05) | 0.88 |
| Xenobiotics | gluconate | -0.09 (-0.18, -0.01) | 0.16 |  | -0.15 (-0.22, -0.07) | 0.004† |  | -0.12 (-0.2, -0.04) | 0.030† |  | -0.01 (-0.09, 0.06) | 0.94 |
|  | 3-phenylpropionate (hydrocinnamate) | 0.06 (-0.03, 0.14) | 0.46 |  | 0.12 (0.05, 0.19) | 0.025† |  | 0.09 (0.01, 0.16) | 0.11 |  | 0.08 (0.01, 0.15) | 0.3 |
|  | p-cresol sulfate | -0.16 (-0.24, -0.08) | 0.006† |  | -0.03 (-0.11, 0.05) | 0.68 |  | -0.09 (-0.17, -0.02) | 0.081 |  | -0.06 (-0.13, 0.01) | 0.53 |
|  | 4-hydroxyhippurate | -0.05 (-0.14, 0.03) | 0.51 |  | -0.13 (-0.2, -0.05) | 0.015† |  | -0.11 (-0.18, -0.04) | 0.033† |  | -0.12 (-0.19, -0.05) | 0.091 |
|  | 4-allylphenol sulfate | 0.13 (0.05, 0.21) | 0.040† |  | 0.09 (0.02, 0.17) | 0.11 |  | 0.09 (0.02, 0.17) | 0.093 |  | 0.06 (-0.02, 0.13) | 0.56 |
|  | ergothioneine | 0.1 (0.01, 0.18) | 0.13 |  | 0.17 (0.08, 0.24) | 0.002† |  | 0.18 (0.11, 0.25) | < 0.001† |  | 0.08 (0.01, 0.15) | 0.3 |
|  | mannonate* | -0.08 (-0.15, 0) | 0.28 |  | -0.14 (-0.22, -0.06) | 0.007† |  | -0.15 (-0.22, -0.07) | 0.003† |  | -0.04 (-0.11, 0.04) | 0.71 |
|  | indolin-2-one | -0.11 (-0.2, -0.02) | 0.074 |  | -0.06 (-0.13, 0.02) | 0.38 |  | -0.12 (-0.2, -0.04) | 0.027† |  | -0.04 (-0.12, 0.04) | 0.73 |
|  | 6-hydroxyindole sulfate | -0.1 (-0.18, -0.01) | 0.13 |  | -0.07 (-0.14, 0.01) | 0.29 |  | -0.11 (-0.19, -0.04) | 0.031† |  | -0.04 (-0.12, 0.04) | 0.74 |
|  | ethyl beta-glucopyranoside | 0 (-0.09, 0.08) | 0.98 |  | 0.11 (0.04, 0.18) | 0.038† |  | 0.07 (-0.01, 0.14) | 0.24 |  | -0.07 (-0.15, 0.01) | 0.44 |

Metabolites listed are those significantly associated with any of the four measures of physical activity by super-pathway using Spearman’s partial correlations, adjusting for age (continuous), sex (men, women), smoking (cotinine detected: yes, no), race (non-White, White), body fat index (continuous). Multiple testing was corrected for using the Benjamini-Hochberg procedure (p-adjust<0.05). Confidence intervals were estimated using bootstrap analysis (N iterations=1000).

Super-pathways were defined by Metabolon. Associations with all serum metabolites are available from **Supplementary Data 1**.

†Statistically significant, p-adjust<0.05.

*Indicates a compound that has not been confirmed based on a standard, but confidence in its identity.

**Indicates a compound for which a standard is not available, but reasonable confidence in its identity, or the information provided.

Abbreviations: aMET= active metabolic equivalent of task, DLW=doubly labelled water, PAL=physical activity level.

Supplementary Table 4:Metabolites with heterogeneous associations with physical activity by sex

| Metabolite | Physical activity measure | Men | | |  | Women | | |  | P-het |
| --- | --- | --- | --- | --- | --- | --- | --- | --- | --- | --- |
|  |  | N | R | P-value |  | N | R | P-value |  |  |
| glycerol 3-phosphate | Steps/d, activPAL | 346 | 0.11 | 0.03 |  | 361 | -0.11 | 0.03 |  | 0.002 |
| 1,5-anhydroglucitol (1,5-AG) | aMET hr/d, ActiGraph | 345 | 0.17 | 0.002 |  | 359 | -0.04 | 0.40 |  | 0.004 |
| X-21834 | aMET hr/d, ActiGraph | 345 | -0.09 | 0.09 |  | 359 | 0.13 | 0.01 |  | 0.003 |
| X-23666 | aMET hr/d, ACT24 | 345 | 0.06 | 0.30 |  | 361 | -0.16 | 0.003 |  | 0.005 |

Correlations estimated using Spearman’s partial correlation, adjusted for age (continuous), smoking (cotinine detected: yes, no), race (non-White, White), body fat index (continuous). Heterogeneity assessed using Cochran’s Q. Statistical significance threshold based on the Benjamini-Hochberg procedure (FDR) in the primary analysis (p-unadjusted<0.005). Full results are available from **Supplementary Data 2**.

Abbreviations: DLW=doubly labelled water, FDR=false discovery rate, aMET=active metabolic equivalent of task, PAL=physical activity level.

Supplementary Table 5: Comparison of physical activity level-metabolite associations with total energy expenditure-metabolite associations, measured by doubly labeled water in the IDATA Study

| Metabolite | Physical activity level | |  | Total energy expenditure | |
| --- | --- | --- | --- | --- | --- |
|  | r | P-value |  | r | P-value |
| 1-methylnicotinamide | 0.15 | < 0.001† |  | 0.17 | < 0.001† |
| N-formylmethionine | -0.14 | 0.001† |  | -0.09 | 0.033 |
| creatinine | -0.19 | < 0.001† |  | -0.21 | < 0.001† |
| glycine | -0.18 | < 0.001† |  | -0.16 | < 0.001† |
| malate | -0.12 | 0.004† |  | -0.11 | 0.013 |
| serine | -0.14 | 0.001† |  | -0.14 | 0.001† |
| uridine | 0.13 | 0.002† |  | 0.11 | 0.011 |
| pseudouridine | -0.16 | < 0.001† |  | -0.13 | 0.002† |
| uracil | 0.13 | 0.002† |  | 0.11 | 0.012 |
| vanillylmandelate (VMA) | -0.12 | 0.004† |  | -0.05 | 0.23 |
| N-acetylneuraminate | -0.14 | < 0.001† |  | -0.1 | 0.017 |
| creatine | 0.12 | 0.004† |  | 0.14 | 0.001† |
| choline | 0.11 | 0.007 |  | 0.14 | 0.001† |
| 3-methoxytyrosine | -0.12 | 0.004† |  | -0.09 | 0.032 |
| stearoyl sphingomyelin (d18:1/18:0) | -0.14 | < 0.001† |  | -0.1 | 0.015 |
| 1-palmityl-GPC (O-16:0) | -0.12 | 0.004† |  | -0.14 | 0.001† |
| N-acetylaspartate (NAA) | -0.22 | < 0.001† |  | -0.21 | < 0.001† |
| homoarginine | 0.13 | 0.003† |  | 0.09 | 0.033 |
| N-acetylglycine | -0.14 | 0.001† |  | -0.12 | 0.004 |
| pro-hydroxy-pro | -0.08 | 0.078 |  | -0.13 | 0.002† |
| N-acetylthreonine | -0.14 | 0.001† |  | -0.12 | 0.003 |
| gamma-glutamylglycine | -0.18 | < 0.001† |  | -0.14 | < 0.001† |
| gamma-glutamylthreonine | -0.13 | 0.003† |  | -0.12 | 0.003 |
| p-cresol sulfate | -0.16 | < 0.001† |  | -0.15 | < 0.001† |
| aconitate [cis or trans] | -0.16 | < 0.001† |  | -0.12 | 0.005 |
| N6-carbamoylthreonyladenosine | -0.13 | 0.002† |  | -0.13 | 0.003 |
| phenylacetylglutamine | -0.13 | 0.002† |  | -0.13 | 0.002† |
| 5,6-dihydrouridine | -0.14 | < 0.001† |  | -0.12 | 0.006 |
| glutamine_degradant* | -0.15 | < 0.001† |  | -0.17 | < 0.001† |
| 1-dihomo-linolenoyl-GPC (20:3n3 or 6)* | 0.14 | < 0.001† |  | 0.14 | 0.001† |
| glutarylcarnitine (C5-DC) | -0.11 | 0.009 |  | -0.13 | 0.002† |
| tiglylcarnitine (C5:1-DC) | 0.12 | 0.004† |  | 0.13 | 0.003 |
| hexadecanedioate (C16-DC) | -0.13 | 0.002† |  | -0.13 | 0.003 |
| hexanoylglutamine | -0.12 | 0.004† |  | -0.08 | 0.047 |
| 4-allylphenol sulfate | 0.13 | 0.003† |  | 0.1 | 0.025 |
| sphingomyelin (d18:1/18:1, d18:2/18:0) | -0.17 | < 0.001† |  | -0.11 | 0.009 |
| palmitoyl sphingomyelin (d18:1/16:0) | -0.15 | < 0.001† |  | -0.1 | 0.019 |
| argininate* | 0.12 | 0.004† |  | 0.13 | 0.003 |
| sphingomyelin (d18:2/16:0, d18:1/16:1)* | -0.17 | < 0.001† |  | -0.1 | 0.016 |
| 1-dihomo-linolenoyl-GPE (20:3n3 or 6)* | 0.16 | < 0.001† |  | 0.17 | < 0.001† |
| sphingomyelin (d18:1/24:1, d18:2/24:0)* | -0.13 | 0.002† |  | -0.1 | 0.018 |
| p-cresol glucuronide* | -0.17 | < 0.001† |  | -0.17 | < 0.001† |
| sphingomyelin (d18:1/20:1, d18:2/20:0)* | -0.15 | < 0.001† |  | -0.09 | 0.028 |
| sphingomyelin (d18:1/20:2, d18:2/20:1, d16:1/22:2)* | -0.14 | < 0.001† |  | -0.1 | 0.015 |
| sphingomyelin (d18:1/22:2, d18:2/22:1, d16:1/24:2)* | -0.17 | < 0.001† |  | -0.13 | 0.002 |
| C-glycosyltryptophan | -0.17 | < 0.001† |  | -0.15 | < 0.001† |
| adipoylcarnitine (C6-DC) | -0.13 | 0.003† |  | -0.07 | 0.08 |
| sphingomyelin (d18:1/17:0, d17:1/18:0, d19:1/16:0) | -0.16 | < 0.001† |  | -0.14 | < 0.001† |
| sphingomyelin (d18:2/24:1, d18:1/24:2)* | -0.19 | < 0.001† |  | -0.15 | < 0.001† |
| 1-palmitoyl-2-dihomo-linolenoyl-GPC (16:0/20:3n3 or 6)* | 0.12 | 0.004† |  | 0.13 | 0.002† |
| 1-(1-enyl-stearoyl)-2-docosahexaenoyl-GPE (P-18:0/22:6)* | -0.13 | 0.002† |  | -0.13 | 0.002 |
| 1-(1-enyl-stearoyl)-2-docosahexaenoyl-GPC (P-18:0/22:6)* | -0.14 | 0.001† |  | -0.15 | < 0.001† |
| 1-stearyl-2-arachidonoyl-GPC (O-18:0/20:4)* | -0.14 | 0.001† |  | -0.14 | 0.001† |
| lactosyl-N-palmitoyl-sphingosine (d18:1/16:0) | -0.13 | 0.002† |  | -0.1 | 0.015 |
| 1-stearoyl-2-dihomo-linolenoyl-GPC (18:0/20:3n3 or 6)* | 0.13 | 0.003† |  | 0.16 | < 0.001† |
| 1-stearyl-GPC (O-18:0)* | -0.14 | < 0.001† |  | -0.16 | < 0.001† |
| 1-stearoyl-2-dihomo-linolenoyl-GPE (18:0/20:3n3 or 6)* | 0.13 | 0.003† |  | 0.12 | 0.003 |
| glycosyl-N-palmitoyl-sphingosine (d18:1/16:0) | -0.15 | < 0.001† |  | -0.12 | 0.006 |
| 1-palmityl-2-palmitoyl-GPC (O-16:0/16:0)* | -0.12 | 0.004† |  | -0.11 | 0.012 |
| sphingomyelin (d18:2/18:1)* | -0.17 | < 0.001† |  | -0.13 | 0.002 |
| sphingomyelin (d18:2/24:2)* | -0.22 | < 0.001† |  | -0.17 | < 0.001† |
| sphingomyelin (d18:2/23:1)* | -0.18 | < 0.001† |  | -0.15 | < 0.001† |
| lignoceroylcarnitine (C24)* | 0.14 | 0.001† |  | 0.1 | 0.018 |
| octadecenedioate (C18:1-DC) | -0.14 | 0.001† |  | -0.14 | < 0.001† |
| octadecadienedioate (C18:2-DC)* | -0.12 | 0.003† |  | -0.15 | < 0.001† |
| hydroxyasparagine** | -0.13 | 0.002† |  | -0.09 | 0.038 |
| glutamine conjugate of C6H10O2 (2)* | -0.13 | 0.003† |  | -0.09 | 0.04 |
| 3-amino-2-piperidone | -0.14 | 0.001† |  | -0.11 | 0.012 |
| hydroxy-N6,N6,N6-trimethyllysine* | -0.14 | < 0.001† |  | -0.11 | 0.008 |
| palmitoyl-sphingosine-phosphoethanolamine (d18:1/16:0) | -0.14 | < 0.001† |  | -0.09 | 0.045 |
| X-12026 | -0.16 | < 0.001† |  | -0.13 | 0.002† |
| X-12112 | 0.12 | 0.005 |  | 0.13 | 0.002† |
| X-12117 | -0.15 | < 0.001† |  | -0.12 | 0.005 |
| X-12306 | 0.17 | < 0.001† |  | 0.14 | 0.001† |
| X-21310 | -0.12 | 0.004† |  | -0.11 | 0.008 |
| X-23666 | -0.17 | < 0.001† |  | -0.17 | < 0.001† |

Correlations estimated using Spearman’s partial correlation, adjusted for age (continuous), sex (men, women), smoking (cotinine detected: yes, no), race (non-White, White). Physical activity level additionally adjusted for body fat index (continuous). Total energy expenditure additionally adjusted for total fat mass (kg, continuous) and fat free mass (kg, continuous).

†Denotes statistical significance based on the Benjamini-Hochberg procedure.

*Indicates a compound that has not been confirmed based on a standard, but confidence in its identity.

Supplementary Table 6: Correlations of physical activity-metabolite associations in IDATA for metabolites included in the exploratory breast cancer analysis

| Metabolite | Physical activity measure | Men | | |  | Women | | |
| --- | --- | --- | --- | --- | --- | --- | --- | --- |
|  |  | N | R | P-value |  | N | R | P-value |
| N-acetylthreonine | PAL, DLW | 276 | -0.17 | 0.005 |  | 280 | -0.09 | 0.13 |
| sphingomyelin (d18:1/20:1, d18:2/20:0)* | PAL, DLW | 276 | -0.15 | 0.02 |  | 280 | -0.16 | 0.009 |
| X-21310 | PAL, DLW | 276 | -0.11 | 0.07 |  | 280 | -0.14 | 0.02 |
| epiandrosterone sulfate | Steps/day, activPAL | 346 | 0.16 | 0.003 |  | 361 | 0.11 | 0.04 |
| beta-hydroxyisovaleroylcarnitine | Steps/day, activPAL | 346 | 0.08 | 0.14 |  | 361 | 0.13 | 0.01 |
| N-acetyltaurine | Steps/day, activPAL | 346 | -0.05 | 0.33 |  | 361 | -0.16 | 0.002 |
| 2-methylbutyrylcarnitine (C5) | aMET hr/d, ActiGraph | 345 | -0.11 | 0.04 |  | 359 | -0.11 | 0.04 |
| andro steroid monosulfate C19H28O6S (1)* | aMET hr/d, ActiGraph | 345 | -0.18 | 0.0008 |  | 359 | -0.13 | 0.02 |
| isovalerylglycine | aMET hr/d, ACT24 | 345 | 0.13 | 0.01 |  | 361 | 0.14 | 0.007 |

Correlations estimated using Spearman’s partial correlation, adjusted for age (continuous), smoking (cotinine detected: yes, no), race (non-White, White), body fat index (continuous) in the IDATA study. Metabolites shown here are those associated with any of the physical activity measure (after correcting for multiple testing using the Benjamini-Hochberg procedure, p-adjust <0.05) in the IDATA Study and nominally associated with breast cancer (p-unadjust<0.05) in PLCO.

*Indicates a compound that has not been confirmed based on a standard, but confidence in its identity.

Abbreviations: aMET=active metabolic equivalent of task, DLW=doubly labelled water, PAL=physical activity level, PLCO=Prostate, Lung, Colorectal and Ovarian Cancer Screening Trial.
